# Artificial Intelligence-Based Automated Interpretation of Images of Electrocardiograms: Development and Multinational Validation of ECG-GPT

**DOI:** 10.1101/2024.02.17.24302976

**Authors:** Akshay Khunte, Veer Sangha, Evangelos K Oikonomou, Lovedeep S Dhingra, Arya Aminorroaya, Andreas Coppi, Sumukh Vasisht Shankar, Elijah Rockers, Bobak J Mortazavi, Deepak L Bhatt, Harlan M Krumholz, Sadeer Al-Kindi, Girish N Nadkarni, Akhil Vaid, Rohan Khera

**Affiliations:** Section of Cardiovascular Medicine, Department of Internal Medicine, Yale School of Medicine, New Haven, CT; NYU Grossman School of Medicine, New York, NY; Department of Engineering Science, Oxford University, Oxford, UK; Center for Outcomes Research and Evaluation, Yale-New Haven Hospital, New Haven, CT; Center for Cardiovascular Computational & Precision Health, Houston Methodist DeBakey Heart & Vascular Center, Houston, TX, USA; Department of Computer Science & Engineering, Texas A&M University, College Station, TX; Mount Sinai Fuster Heart Hospital, Icahn School of Medicine at Mount Sinai, New York, NY; Department of Health Policy and Management, Yale School of Public Health, New Haven, CT; The Charles Bronfman Institute for Personalized Medicine, Icahn School of Medicine at Mount Sinai, New York, NY, USA; The Division of Data Driven and Digital Medicine, Department of Medicine, Icahn School of Medicine at Mount Sinai, New York, NY, USA; Section of Health Informatics, Department of Biostatistics, Yale School of Public Health, New Haven, CT

## Abstract

**Background:** Timely and accurate assessment of electrocardiograms (ECGs) is crucial for diagnosing, triaging, and clinically managing patients. Current workflows rely on computerized ECG interpretation tools built into ECG signal acquisition systems, which use rule-based algorithms that are unreliable and frequently not available in low-resource settings. We developed and validated a format-independent vision encoder-decoder model – ECG-GPT – that can generate free-text, expert-level interpretations directly from 12-lead ECG images.

**Methods:** Using 12-lead ECGs and their corresponding diagnosis statements collected at the Yale-New Haven Health System (YNHHS) between 2000 and 2022, we developed a vision-text transformer model to generate interpretation statements from images of ECGs. Using structured clinical assessment, semantic similarity, and conventional natural language generation metrics, we validated ECG-GPT across 7 geographically distinct health settings. These include (1) 3 large and diverse US health systems, (2) consecutive ECGs from a central reading system in Minas Gerais, Brazil, (3) the prospective cohort study, UK Biobank, (4) a Germany-based, publicly available repository, PTB-XL, and (5) a community hospital in Missouri.

**Results:** Overall, 2.9 million ECGs were used for model development. The model performed well in clinical assessment across 26 extracted labels: for atrial fibrillation, sinus tachycardia, sinus bradycardia, premature atrial contractions, and premature ventricular contractions, AUROCs and AUPRCs ranged from 0.80-0.95 and 0.50-0.86, respectively. For left bundle branch block, right bundle branch block, first degree atrioventricular block, left anterior fascicular block, and left posterior fascicular block, AUROCs and AUPRCs ranged from 0.88-0.96 and 0.23-0.86, respectively. Across all 26 conditions, diagnostic accuracy ranged between 0.93-0.99. ECG-GPT identified the full context of the diagnosis statements with allied conditions. It had a median pairwise cosine similarity of 0.90 (IQR 0.83-0.97), significantly greater than the median baseline similarity of 0.73 (IQR 0.67-0.78, p<0.001). This separation between median pairwise and baseline similarity remained consistent across all 26 condition-specific subsets. The results were comparable across external validation sites.

**Conclusions:** We developed and extensively validated a vision encoder-decoder model that generates expert-level interpretations from ECG images. This represents a scalable and accessible strategy for automated ECG analysis, especially in low-resource settings.

**CLINICAL PERSPECTIVE:** *What is New?:*

- ECG-GPT is a vision encoder-decoder model capable of generating full-text ECG interpretations directly from ECG images, regardless of layout or format.
- The model was trained on over 2.7 million ECGs and externally validated across 3.8 million additional ECGs from demographically and geographically diverse populations.

*What are the clinical implications?:*

- ECG-GPT enables automated, expert-level ECG interpretation directly from images, eliminating the need for signal data or device integration.
- The model demonstrates consistent performance across diverse patient populations, ECG formats, and care settings.
- This scalable, image-based approach may expand access to accurate ECG interpretation in low-resource settings.

## INTRODUCTION

Electrocardiography (ECG) is a widely available, first-line, noninvasive tool for diagnosing, triaging, and managing cardiovascular disease.^1^ Traditional workflows often rely on computerized ECG interpretation algorithms to generate preliminary reads, which, despite limited accuracy,^2,3^ provide diagnostic support and enable faster triage for high-risk conditions.^4^ Such algorithms, however, are often proprietary, require raw signal data, and have substantial variability in accuracy,^4,5^ with the potential to lead to patient harm.^6,7^ This makes computerized pre-reads inaccessible to clinicians in rural and low-resource settings, a disparity further exacerbated by the lower availability of expert-level readers.^8,9^ This lack of reliable and interoperable automated system-generated ECG reports in many low-resource settings globally highlights the need for an accurate, easily accessible ECG interpretation tool.

This is particularly pronounced in community health centers in rural areas of low- to middle-income countries, where junior health professionals and community health workers provide preventive and basic care, including the interpretation of ECGs. Further management frequently relies on telecommunications with a limited number of experts available to provide such consultation.^10^ Moreover, the accuracy of ECG interpretations by physicians is variable even after educational interventions, with less than 75% accuracy reported for even cardiologists.^11^ Therefore, there is an unmet need for accessible, reliable, and accurate tools to provide expert-level ECG interpretations.

Though recent advances in deep learning enable accurate classification of specific ECG abnormalities,^12–15^ they are generally limited to a select number of commonly encountered abnormalities and do not address less common conduction and rhythm disorders or variations within common rhythms. However, even these less common abnormalities may have significant therapeutic and prognostic implications despite their low prevalence. Additionally, while our prior work has demonstrated that diagnostic models can directly identify information from ECG images,^15,16^ most traditional signal-based models, like computerized interpretation algorithms, predominantly rely on raw signal data, limiting their scalability to the point-of-care and across low-resource settings. The development of models exclusively for images is challenged by variations in the layout of the leads, labeling of the leads, structure and design of the graphed background, and the quality of the image acquisition.

In this study, we report the development of ECG-GPT (**Figure 1**), a novel vision-text transformer model capable of generating diagnostic reports from ECG images regardless of the layout, trained against the full breadth of expert-verified ECG interpretations across 2.7 million 12-lead ECGs collected over 21 years in a large US-based hospital system. ECG-GPT can be accessed as a web-based application that can receive ECG images across formats and layouts as the only input and generate diagnostic reports (demonstration hosted at https://www.cards-lab.org/ecg-gpt). We pursued broad multinational validation in 3.8 million ECGs across temporally and geographically distinct datasets, including two distinct US-based major referral hospital systems, a Brazil-based large telehealth network, a UK-based prospective cohort study, a publicly available ECG dataset from Germany, and a rural US-based community hospital.

**Figure 1.**
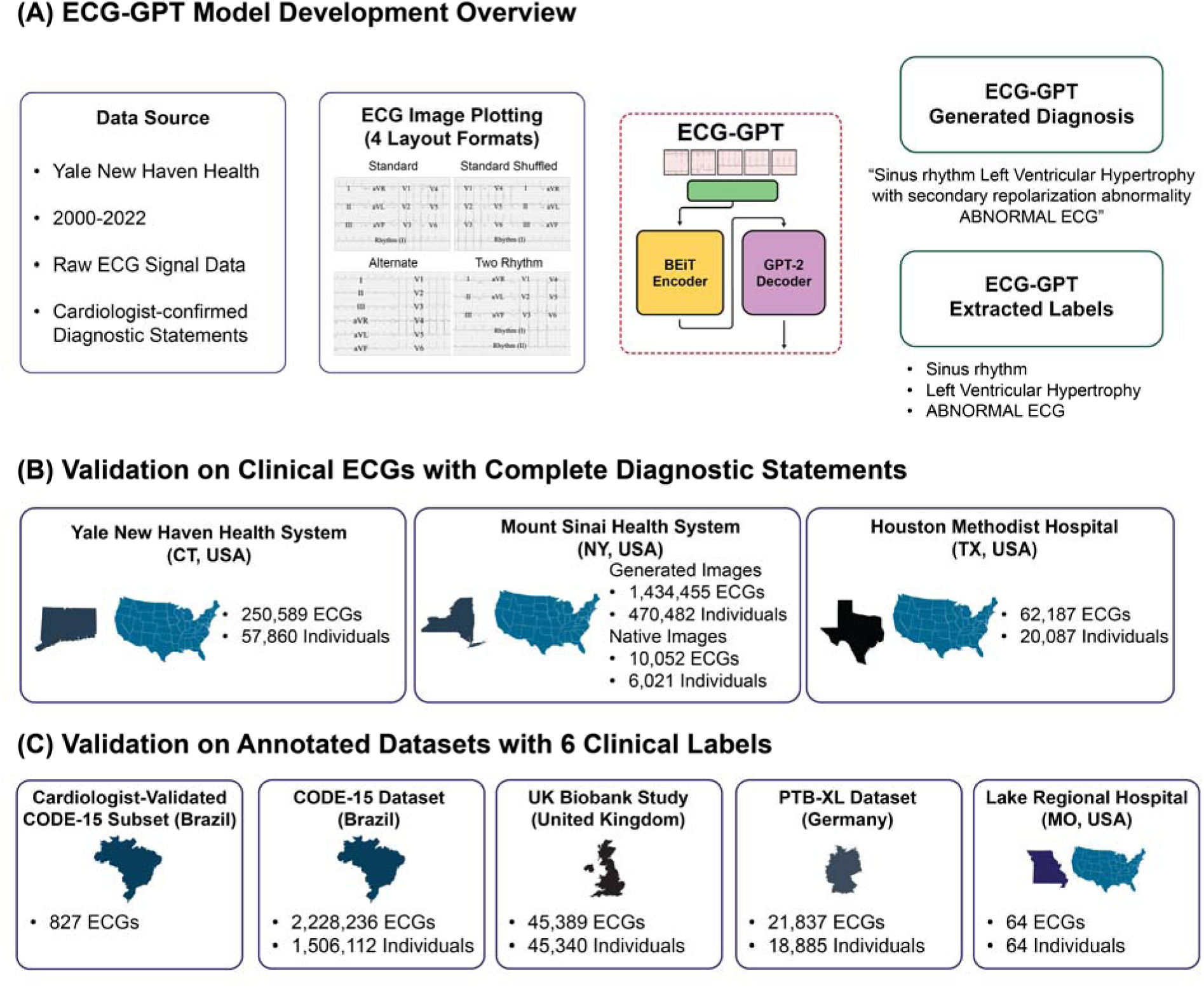
Overview of ECG-GPT’s development and evaluation. Abbreviations: ECG, electrocardiogram; BEiT, Bidirectional Encoder representation from Image Transformers; GPT-2, Generative Pretrained Transformer-2.

## METHODS

The study was reviewed by the Yale Institutional Review Board, which approved the study protocol and waived the need for informed consent as the study represents secondary analysis of existing data. The development dataset from Yale and the validation datasets from Mount Sinai, Houston Methodist Hospital, and Lake Regional Hospital are not publicly available, given the stipulations of the relevant Institutional Review Boards. The external datasets from Brazil, UK Biobank, and PTB-XL are available directly from the respective groups and are outside the purview of the authors. The model is publicly available online as a web-based application for research use at https://www.cards-lab.org/ecg-gpt.

### Data Source for Model Development

Raw voltage data were collected for all 12-lead ECGs with corresponding diagnosis statements obtained at the Yale New Haven Health System (YNHHS) during 2000-2021. Each ECG was recorded with a sampling frequency of 500 Hz using Philips PageWriter and GE MAC machines. The subset of these ECGs with continuous recording across all 12 leads was then split at the patient level into training, validation, and test sets (85%, 5%, 10%). For the test set, we restricted to ECGs with no marked abnormalities and abnormal ECGs with confirmed reports certified by a cardiologist. In the training and validation sets, we chose to include abnormal, unconfirmed ECGs collected in the emergency setting, since these are interpreted at the point-of-care, and may not be listed as a confirmed read in the system. Finally, in the training set, a random set of ECGs with no marked abnormalities were removed to match the observed prevalence of such ECGs in the original cohort (**Figure S1**).

### Signal Preprocessing and ECG Image Generation

First, all ECGs that did not contain 10 seconds of continuous recording across all 12 leads were excluded. To enable the generation of ECG images like those used in clinical settings, we further preprocessed the signal before plotting. For this, we subtracted a one-second median filter from the original raw voltage for each lead of 10-second ECGs to remove baseline wander, mirroring the approach undertaken by ECG machines before printing clinical ECGs available to and interpreted by physicians. All ECGs obtained in the YNHHS were plotted at their original sampling rate of 500 Hz. ECGs used for external validation, which were recorded at sampling frequencies between 300-600 Hz, were down sampled to 300 Hz before plotting, as described previously.^15^ While the ECGs in the development set were recorded using GE MAC and Philips PageWriter machines, those in the external validation were frequently only GE MAC models, even though the specific models used varied within individual validation datasets.

The preprocessed ECG signals were transformed into ECG images at 100 dots per inch (DPI) using the python library ecg-plot.^17^ We employed multiple strategies to ensure the robustness of the model. First, we converted each signal waveform to multiple images using four different layouts of the leads to account for different schemes of real-world ECGs. To ensure the model is resilient to these other formats, it was trained using all these variations of ECG images, an approach we previously developed and validated.^15^ The four formats used in model development (**Figure 1**) included (1) The standard printed ECG format in the US with lead I as the rhythm strip. This format consists of four 2.5-second sequential columns, each containing a 2.5-second strip from three leads. (2) The same as the standard US format that includes an additional rhythm strip from lead II. (3) An alternate format with no rhythm strip comprising two 5-second columns. The first column represents a simultaneous recording from limb leads, while the second column represents a simultaneous recording from precordial leads. (4) A shuffled format in which precordial leads are recorded in the first two columns and limb leads are represented in the last two columns.

Second, the conversion of ECG signals to images was done independently of the model development to ensure the model remains agnostic to preprocessing steps from ECG signals to images. Third, all images were rotated by a random amount between −10 to 10 degrees prior to training. Finally, we used Python Image Library (PIL v9.2.0) to convert all ECG images to greyscale and down-sample them to the required size for input into the model regardless of their initial resolution. Training the model using plotted ECG images instead of raw signal data translates ECG image inference as a computer vision application rather than a signal extraction or processing task. ECG-GPT leverages the entire ECG representation during inference, incorporating background gridlines and lead labels. This approach, combined with the varied formats used during training, enables generating interpretations directly from the ECG images available to end-user clinicians.

### Image Standardization for Model Inference

We have implemented a previously validated approach to standardize model inputs.^16^ Inputs are limited to 12-lead ECG tracings, which are vertically oriented, minimally rotated, have a uniform background, and do not have peripheral annotations. Additionally, to mitigate the effects of noise, a two-step preprocessing approach is applied to each image: first, the image is straightened and cropped to correct for rotations and to remove any elements outside of the ECG tracing, including any potential identifiers or interpretation summary. Second, the algorithm scales the brightness and contrast of the ECGs to the mean values of the development population before generating model predictions (**Figure S2**). ECGs with deviations in brightness and contrast 50% greater or lower than those seen in the development set are flagged as requiring the image to be recaptured in better quality before inference (**Figure S3**).

### Diagnostic Statement Preprocessing

Cardiologist-confirmed diagnosis statements were preprocessed to remove all identifying information using a rule-based approach. A string search was performed for all diagnostic statements to identify and remove names, references to previous ECGs, all dates and times, and punctuation. The Python PyEnchant package was then used to generate a list of all abbreviations present in the processed diagnosis statements.^18^ A pair of clinical experts manually generated a dictionary containing each abbreviation and its expanded form for the 100 most common abbreviations (**Table S1**). Each instance of these abbreviations in the diagnosis statement was then replaced with its expansion. Additionally, the most common misspellings and all synonyms for a condition were replaced with a single term. This processed diagnosis statement was then used for model development and evaluation (**Figure S4**).

### Model Development

We built a custom Vision Encoder-Decoder model using the HuggingFace framework.^19^ For the image encoder, we selected a BEiT transformer model, pretrained on the ImageNet dataset,^20^ due to its robust performance on the ImageNet-1K benchmark, relatively few trainable parameters compared to other state-of-the-art models, and the 384×384 pixel input size.^21^ We selected the base-size version of the model, with ∼84 million trainable parameters, which takes a linearly embedded sequence of 16×16 pixel patches as the input.

We selected the Generative Pretrained Transfomer-2 (GPT-2) transformer for the text decoder, initially developed for prompt-based text generation. This architecture, which contains ∼124 million parameters, decoded the lower-dimensional representations generated from images by the BEiT encoder into ECG interpretation statements with a maximum output token length of 48 tokens. To better tailor the model outputs to the text found in ECG diagnosis statements and to reduce the number of tokens needed, thus lowering the computational costs of using the model, we first fine-tuned the GPT-2 tokenizer on the ECG diagnosis statements used to train the model. In addition to the lightweight nature of GPT-2 relative to other large language models capable of text generation, the extensive pretraining of the GPT-2 decoder in combination with the fine-tuned tokenizer enabled direct integration into the vision-text transformer architecture without further fine-tuning.

Overall, the composed Vision Encoder-Decoder model, consisting of the BEiT encoder and GPT-2 decoder, has over 239 million trainable parameters (**Figure 2**). The model was trained on a generated image dataset at a learning rate of 5 x 10^-5^ for 20 epochs. The model randomly selected a format each time an ECG was loaded during training to reduce overfitting. We used the Adam optimizer, a minibatch size of 14, and a cross-entropy loss function to minimize the error between the GPT-2 output and the tokenized reference diagnostic statements to train the model.^22^ Model development used the HuggingFace Transformers 4.28.1 framework with PyTorch 2.0.0 and Python 3.11.3 on eight NVIDIA A100 80GB graphics processing units.

**Figure 2.**
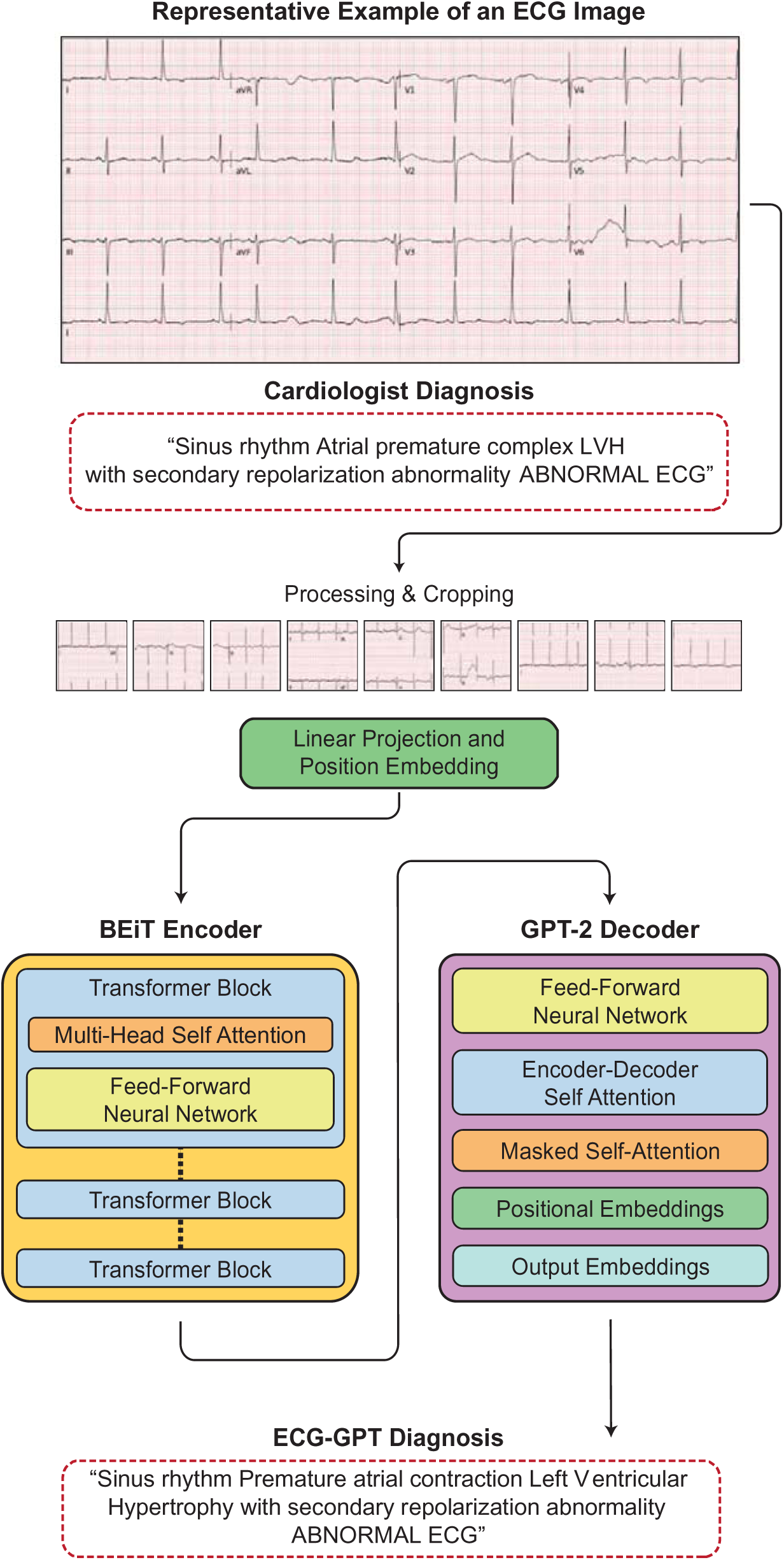
**Vision Encoder-Decoder Model Architecture and Sample ECG-GPT Output Interpretation Statement**. Abbreviations: ECG, electrocardiogram; BEiT, Bidirectional Encoder representation from Image Transformers; GPT-2, Generative Pretrained Transformer-2.

### External Validation

We pursued validation on five generated ECG image datasets and three native ECG image datasets acquired outside the YNHHS. These included A) one generated image dataset and one distinct native image dataset from Mount Sinai Health System (MSHS) in New York, NY, B) a native image dataset from Houston Methodist Hospital (HMH) in Houston, TX, C) a native image dataset from Lake Regional Hospital (LRH) in Osage Beach, MO, and D) images generated from four publicly available ECG signal datasets.

Both sets from MSHS and the set from HMH contained corresponding diagnosis statements. The first set from MSHS included 1,434,455 ECGs collected from 2013 to 2023 that were then plotted in the standard image format used in training. The second included 10,052 distinct, native ECG images collected prospectively from May to July 2024. The set from HMH included 62,876 native ECG images collected from 2018 to 2024. A federated approach was implemented to enable external validation within the MSHS and HMH. Both the model and all semantic and clinical assessment code were containerized using Docker and securely deployed within the infrastructure of each hospital. It accepted file paths as input, ensuring that patient data remained within the hospital’s system without the need for external data sharing. This approach facilitated accurate prediction generation while safeguarding patient privacy. Notably, the lightweight design of the model enables GPU-less inference on devices with at least 16GB of RAM. This, combined with the containerized approach, allows for efficient local deployment on a wide array of machines, enhancing accessibility in settings with limited internet connectivity.

The set from LRH included 64 native ECG images with diagnostic labels for six rhythm and conduction disorders, including atrial fibrillation (AF), sinus tachycardia (ST), sinus bradycardia (SB), left bundle branch block (LBBB), right bundle branch block (RBBB), and first degree atrioventricular block (1dAVb). The set included 8-10 ECGs with each of these labels and ECGs labeled as normal. Though the layout of these ECGs was similar to the standard layout used for model development, there were multiple key distinctions. First, the V1 lead, instead of lead I, was the rhythm lead. Second, the signal was black, as opposed to blue, and vertical lines were separating the leads. Additionally, there were variations in background and grid color, as well as in the position and font of the lead label.

Validation was also pursued in images generated from four geographically distinct, open-source ECG signal datasets labeled with the same six rhythm and conduction disorders as the set from LRH. First, we obtained 45,389 ECGs from the UK Biobank, under research application #71033. The UK Biobank represents the largest population-based cohort of 502,468 people in the United Kingdom with protocolized imaging, laboratory testing, and linked electronic health records. We also used a set of 2,228,236 ECGs from the CODE15 study dataset, a set of ECG recordings previously used for both signal- and image-based multilabel ECG classification models.^12,15,23^ As the primary CODE15 dataset consists of ECGs collected and annotated for six rhythm and conduction disorders by individual clinicians during routine care, we also deployed the model in a secondary cardiologist-validated dataset. This dataset contained 827 additional ECGs collected in the Telehealth Network of Minas Gerais between April and September 2018.^12,15^ For each of these ECGs, annotations for the six rhythm and conduction disorders were made by two independent cardiologists following criteria from the American Heart Association,^24^ with disagreements resolved by a third cardiologist. Finally, the model performance was assessed in PTB-XL, a previously described dataset of ECGs.^25^ This dataset contains 21,837 10-second, 12-lead recordings collected at 500 Hz from 18,885 patients in Germany between 1989 and 1996. The records for each ECG, including diagnostic, form, and rhythm statements, were used to extract labels for the same set of rhythm and conduction disorders.

### Model Evaluation

We evaluated ECG-GPT using three distinct strategies: (1) semantic similarity to assess the model’s ability to capture the full clinical context of reference interpretations, (2) syntactic similarity to quantify the overlap between reference and generated statements, and (3) structured label assessment to evaluate the model’s accuracy for specific clinical labels extracted from the statements. Clinical labels for 26 conditions, spanning key rhythm and conduction disorders selected by two cardiologists, were extracted from each reference and model-generated statement using a standardized string search approach. For each condition, basic string search was performed to extract each label using a set of strings (**Table S2**). A condition was flagged as positive if the statement contained a full match for any string in the set. ECGs were flagged as negative if there was no match or if the match was preceded by a negation, including “no” or “without”.

First, for semantic similarity, we fine-tuned a lightweight DistilBERT model,^26,27^ pretrained on a large corpus of electronic health record notes,^28,29^ in the same set of cardiologist-confirmed diagnosis statements used to train the vision-text transformer model. The training mirrored the standard approach for training masked language models, with a chunk size of 128 tokens and a masking probability of 0.15. The model was trained at a 2 x 10^-5^ learning rate until validation loss did not improve for three consecutive epochs. We used the Adam optimizer, a minibatch size of 16, and a cross-entropy loss function to minimize the error between the masked and generated tokens to train the model.^22^ Model development used the HuggingFace Transformers 4.28.1 framework with Torch 2.0.0 and Python 3.11.3 on four RTX 3090 graphics processing units.

After fine-tuning, we deployed the masked language model in the held-out test set and the MSHS external validation set to generate 768-dimensional embeddings for each reference and model-generated statement. We used an identical federated approach to the model-generated statements to generate embeddings for the statements within the MSHS and HMH. Pairwise similarity was computed as the median cosine similarity between the embeddings for each reference statement and its paired model-generated statement. Baseline similarity was computed as the median cosine similarity between the embeddings for 100,000 random pairings of reference and model-generated statements. Of note, there are no benchmarks for semantic similarity, as its interpretation varies based on the specific task and data context. Cosine similarity values range from 0 to 1, with higher values indicating higher similarity between two statements.

To assess the model’s ability to diagnose specific conditions within their complete clinical context, we created subsets for each of the 26 diagnostic labels identified from the reference statements, consisting of all ECGs marked positive for that condition. Pairwise and baseline similarity were computed identically to the approach used for the complete datasets. For subsets too small to generate 100,000 random pairings, the baseline similarity was reported as the median cosine similarity between embeddings for all possible reference and model-generated statement pairings within the subset.

Next, we used four conventional NLG metrics to assess the syntactic similarity between the original diagnosis reports and generated text. ROUGE and BLEU, which range from 0 to 1, evaluate recall and precision, respectively, for the overlap of n-grams between generated and reference statements, providing insight into content overlap and coherence.^30,31^ METEOR, which ranges from 0 to 1, incorporates both syntactic and semantic similarity by aligning word stems and synonyms, enabling an evaluation of content relevance in addition to word overlap.^32^ Finally, CIDEr, which ranges from 0 to 5, measures consensus between generated text and reference summaries through similarity to human consensus, enhancing evaluation robustness across various linguistic styles.^33^ Collectively, these metrics offer a comprehensive assessment of the syntactic similarity between original diagnosis statements and model-generated statements. Each of these metrics was deployed in both the internal held-out test set using the HuggingFace Evaluate package for computing ROUGE, BLEU, and METEOR scores and the COCO Caption Evaluation package for computing CIDEr scores, respectively.^34,35^

Finally, we also implemented a secondary analysis of model performance using the 26 extracted labels. For each reference and model-generated statement, we computed agreement between the statements for each of the 26 rhythm and conduction disorders. For the open-source datasets, we compared the corresponding reference labels for AF, ST, SB, RBBB, LBBB, and 1dAVb to the labels extracted from the model-generated statements for these ECGs. We used multiple metrics to comprehensively evaluate ECG-GPT’s performance on structured labels, including the area under the receiver operating characteristic (AUROC) to measure model discrimination between classes. 95% confidence intervals for AUROC were calculated using DeLong’s algorithm.^36,37^ We also assessed the area under precision-recall curve (AUPRC), which is especially informative for imbalanced classes as it emphasizes correct identification of the positive class. We employed the bootstrap resampling method to estimate confidence intervals for AUPRC.^38^ Additionally, we evaluated accuracy, sensitivity, specificity, F1 score, positive predictive value (PPV), and negative predictive value (NPV) across conditions to provide a thorough assessment of the model’s performance.

### Statistical Analysis

Summary statistics are presented as counts and percentages for categorical elements and median and interquartile range (IQR) for continuous elements. A paired t-test was used to compute the probability of overlap between the pairwise and baseline cosine similarity of reference and model-generated statements. All analyses were performed using Python 3.11.3, and the significance level was set at an alpha of 0.05.

## RESULTS

### Study Population

We developed ECG-GPT in a set of 2,888,384 12-lead ECG recordings with accompanying cardiologist-confirmed diagnosis statements performed on 601,616 unique patients at the Yale-New Haven Health System (YNHHS) between 2000 and 2022. These ECGs reflected a wide distribution of demographics, with a mean age of 63.2 (SD 18.0) years at the time of the ECG. In the development population, 1,393,683 (48.2%) of the ECGs were from women, 1,904,662 (65.9%) ECGs were from non-Hispanic White, 437,244 (15.1%) non-Hispanic Black, 298,490 (10.3%) Hispanic, 34,855 (1.2%) non-Hispanic Asian, and 213,133 (7.4%) from patients from other racial backgrounds (**Table S3**), reflecting broad and diverse representation. In a period when direct linkage to hospital admission records data was possible (with the digitization of health records, starting 2013), 37.5%, 13.5%, and 49.0% of ECGs were performed in the inpatient, emergency, and outpatient settings, respectively.

After processing with a rule-based approach, the diagnosis statements corresponding to each ECG had a median length of 81 characters (IQR 51, 120) and a median length of 12 tokens (IQR 8, 17) after tokenization. In the development cohort, most ECGs reported sinus rhythm (1,935,574, 67.0%), followed by left atrial enlargement (359,747, 12.5%), left ventricular hypertrophy (342,340, 11.9%), and atrial fibrillation (AF) (281,961, 9.8%). A total of 265,640 (9.2%) and 126,757 (4.4%) ECGs reported sinus tachycardia (ST) and sinus bradycardia (SB), respectively. Right bundle branch block (RBBB) and left bundle branch block (LBBB) were present in 195,474 (6.8%) and 94,928 (3.3%) ECGs, respectively, and 211,836 (7.33%) ECGs reported first degree atrioventricular block (1dAVb). Though the presence of multiple rhythms over a 10-second duration would be unusual, if the patient had a series of conduction and rhythm disorders all present together (such as AF with premature ventricular contractions and bundle branch blocks), all would be included in the original diagnosis statement and thus the extracted labels. Accordingly, 104,451 (3.6%) ECGs featured multiple key rhythm disorders, including AF, ST, SB, premature atrial contractions (PACs), and premature ventricular contractions (PVCs). The proportion of ECGs with the 26 conditions, spanning rhythm and conduction disorders extracted from the diagnosis statements is listed in **Table S3**.

### Semantic Similarity

We used a fine-tuned DistilBERT language model to generate embeddings for each reference and model-generated statements for each ECG in the 250,589 ECGs in the internal held-out test set, obtained from 57,860 patients who had not contributed any data to the training or validation set (**Table S3**). The median cosine similarity between the embeddings for reference statements and their paired model-generated statements was 0.90 (IQR 0.83-0.97). This result was significantly higher than the median cosine similarity between 100,000 randomly selected combinations of reference and model-generated statements (0.73 (IQR 0.67-0.78, p<0.001).

In a secondary analysis to assess the model’s ability to capture individual conditions within their full clinical context, the median pairwise cosine similarity was significantly greater than the respective median random cosine similarity across all 26 subsets (**Table 1**). Across all 26 conditions, pairwise and baseline similarities ranged from 0.84-0.97 and 0.72-0.84, respectively. For key rhythm disorders – AF, ST, SB, premature atrial complexes (PACs), and premature ventricular complexes (PVCs) – pairwise and random similarities ranged from 0.91-0.93 and 0.77-0.81, respectively. For conduction abnormalities – LBBB, RBBB, 1dAVb, left anterior fascicular block (LAFB), and left posterior fascicular block (LPFB) – pairwise and random similarities ranged from 0.91-0.95 and 0.79-0.84, respectively.

**Table 1.** Pairwise and baseline similarity between reference and model-generated ECG interpretation statements in the held-out test set. Abbreviations: AF, atrial fibrillation; ST, sinus tachycardia; SB, sinus bradycardia; LBBB, left bundle branch block; RBBB, right bundle branch block; LAFB, left anterior fascicular block; LPFB, left posterior fascicular block; SVT, supraventricular tachycardia; PAC, premature atrial complexes; PVC, premature ventricular complexes; LAE, left atrial enlargement; RAE, right atrial enlargement; LVH, left ventricular hypertrophy; RVH, right ventricular hypertrophy; MI, myocardial infarction; AVb, atrioventricular block; WPW, Wolff-Parkinson-White syndrome; Repol, repolarization. *Acute MI includes ST Elevation MI (STEMI).

| Labels | Pairwise Similarity | Baseline Similarity | P-Value |
| --- | --- | --- | --- |
| Overall | 0.896 (0.827-0.970) | 0.727 (0.671-0.783) | <0.001 |
| Sinus Rhythm | 0.884 (0.815-0.968) | 0.746 (0.691-0.800) | <0.001 |
| AF | 0.914 (0.859-0.968) | 0.794 (0.748-0.839) | <0.001 |
| Atrial Flutter | 0.909 (0.844-0.961) | 0.782 (0.740-0.839) | <0.001 |
| ST | 0.913 (0.849-0.975) | 0.788 (0.739-0.837) | <0.001 |
| SB | 0.945 (0.871-1.000) | 0.807 (0.755-0.860) | <0.001 |
| Sinus Arrhythmia | 0.969 (0.911-1.000) | 0.819 (0.743-0.884) | <0.001 |
| LBBB | 0.929 (0.846-1.000) | 0.824 (0.768-0.880) | <0.001 |
| RBBB | 0.939 (0.885-0.994) | 0.826 (0.778-0.874) | <0.001 |
| LAFB | 0.931 (0.875-0.975) | 0.803 (0.756-0.850) | <0.001 |
| LPFB | 0.949 (0.897-0.989) | 0.842 (0.787-0.891) | <0.001 |
| SVT | 0.899 (0.828-0.955) | 0.778 (0.726-0.828) | <0.001 |
| PAC | 0.908 (0.848-0.956) | 0.769 (0.715-0.821) | <0.001 |
| PVC | 0.927 (0.874-0.969) | 0.784 (0.736-0.830) | <0.001 |
| LAE | 0.914 (0.841-0.968) | 0.766 (0.713-0.822) | <0.001 |
| RAE | 0.920 (0.866-0.964) | 0.770 (0.718-0.823) | <0.001 |
| LVH | 0.935 (0.879-0.979) | 0.799 (0.745-0.853) | <0.001 |
| RVH | 0.954 (0.882-0.987) | 0.794 (0.736-0.858) | <0.001 |
| Low Voltage | 0.914 (0.845-0.968) | 0.756 (0.698-0.816) | <0.001 |
| Left Axis Deviation | 0.913 (0.851-0.969) | 0.760 (0.712-0.810) | <0.001 |
| Acute MI | 0.902 (0.840-0.958) | 0.764 (0.717-0.819) | <0.001 |
| Lead Reversal | 0.923 (0.844-0.969) | 0.816 (0.732-0.889) | <0.001 |
| 1st degree AVb | 0.913 (0.849-0.967) | 0.787 (0.736-0.840) | <0.001 |
| 2nd degree AVb | 0.898 (0.837-0.948) | 0.815 (0.759-0.865) | <0.001 |
| 3rd degree AVb | 0.870 (0.801-0.929) | 0.753 (0.690-0.807) | <0.001 |
| WPW | 0.837 (0.719-0.935) | 0.724 (0.670-0.838) | <0.001 |
| Repol Abnormality | 0.922 (0.858-0.971) | 0.780 (0.726-0.840) | <0.001 |

### Syntactic Similarity

For NLG metrics, ECG-GPT matches or outperforms most state-of-the-art medical image captioning models.^39–41^ We report scores of 0.663 and 0.655 for ROUGE-1 and ROUGE-L, respectively. For BLEU scores we report scores ranging from 0.595 for BLEU-1 to 0.419 for BLEU-4. We also report a METEOR score of 0.693 and a CIDEr score of 3.32 (**Table S4**).

### Structured Label Assessment

Model performance in the held-out test set for each of the 26 rhythm and conduction disorders, including accuracy, positive and negative predictive values, specificity, sensitivity, AUROC, AUPRC, and F1 scores, are recorded in **Table 2**. For AF, ST, SB, PACs, and PVCs, AUROCs and AUPRCs ranged from 0.80-0.95 and 0.50-0.86, respectively. For LBBB, RBBB, 1dAVb, LAFB, and LPFB, the AUROCs and AUPRCs ranged from 0.88-0.96 and 0.23-0.86, respectively. Across all 26 conditions, diagnostic accuracy ranged between 0.93-0.99.

**Table 2.** Clinical assessment of model-generated ECG interpretation statements in the held-out test set. Abbreviations: PPV, Positive Predictive Value; NPV, Negative Predictive Value; AUROC, area under the receiver operator characteristic; AUPRC, area under precision-recall curve; AF, atrial fibrillation; ST, sinus tachycardia; SB, sinus bradycardia; LBBB, left bundle branch block; RBBB, right bundle branch block; LAFB, left anterior fascicular block; LPFB, left posterior fascicular block; SVT, supraventricular tachycardia; PAC, premature atrial complexes; PVC, premature ventricular complexes; LAE, left atrial enlargement; RAE, right atrial enlargement; LVH, left ventricular hypertrophy; RVH, right ventricular hypertrophy; MI, myocardial infarction; AVb, atrioventricular block; WPW, Wolff-Parkinson-White syndrome; Repol, repolarization. *Acute MI includes ST Elevation MI (STEMI).

| Labels | Accuracy | PPV | NPV | Specificity | Sensitivity | AUROC | AUPRC | F1 |
| --- | --- | --- | --- | --- | --- | --- | --- | --- |
| Sinus Rhythm | 0.950 | 0.971 | 0.913 | 0.944 | 0.954 | 0.949 (0.948-0.95) | 0.956 (0.955-0.957) | 0.962 |
| AF | 0.970 | 0.853 | 0.984 | 0.983 | 0.862 | 0.922 (0.92-0.925) | 0.75 (0.745-0.755) | 0.858 |
| Atrial Flutter | 0.968 | 0.381 | 0.997 | 0.970 | 0.868 | 0.919 (0.914-0.923) | 0.333 (0.324-0.342) | 0.529 |
| ST | 0.988 | 0.936 | 0.992 | 0.994 | 0.912 | 0.953 (0.951-0.955) | 0.86 (0.856-0.865) | 0.924 |
| SB | 0.988 | 0.902 | 0.992 | 0.995 | 0.867 | 0.931 (0.928-0.934) | 0.79 (0.783-0.797) | 0.885 |
| Sinus Arrhythmia | 0.963 | 0.395 | 0.998 | 0.964 | 0.922 | 0.943 (0.94-0.947) | 0.366 (0.358-0.373) | 0.553 |
| LBBB | 0.990 | 0.853 | 0.995 | 0.995 | 0.855 | 0.925 (0.921-0.929) | 0.735 (0.725-0.744) | 0.854 |
| RBBB | 0.989 | 0.909 | 0.995 | 0.993 | 0.935 | 0.964 (0.962-0.965) | 0.854 (0.849-0.859) | 0.922 |
| LAFB | 0.971 | 0.701 | 0.989 | 0.980 | 0.808 | 0.894 (0.891-0.898) | 0.577 (0.568-0.583) | 0.751 |
| LPFB | 0.987 | 0.271 | 0.999 | 0.988 | 0.861 | 0.924 (0.915-0.933) | 0.234 (0.222-0.248) | 0.412 |
| SVT | 0.994 | 0.399 | 0.999 | 0.996 | 0.698 | 0.847 (0.833-0.861) | 0.28 (0.259-0.303) | 0.508 |
| PAC | 0.967 | 0.771 | 0.976 | 0.989 | 0.616 | 0.802 (0.798-0.806) | 0.498 (0.49-0.506) | 0.685 |
| PVC | 0.973 | 0.799 | 0.988 | 0.983 | 0.848 | 0.915 (0.913-0.918) | 0.688 (0.681-0.694) | 0.823 |
| LAE | 0.927 | 0.679 | 0.961 | 0.956 | 0.706 | 0.831 (0.828-0.834) | 0.513 (0.508-0.519) | 0.692 |
| RAE | 0.989 | 0.251 | 0.999 | 0.990 | 0.764 | 0.877 (0.864-0.889) | 0.192 (0.178-0.207) | 0.377 |
| LVH | 0.949 | 0.729 | 0.983 | 0.960 | 0.864 | 0.912 (0.910-0.914) | 0.645 (0.641-0.651) | 0.791 |
| RVH | 0.927 | 0.118 | 0.998 | 0.928 | 0.822 | 0.875 (0.868-0.882) | 0.099 (0.095-0.104) | 0.206 |
| Low Voltage | 0.945 | 0.660 | 0.977 | 0.963 | 0.762 | 0.862 (0.86-0.865) | 0.524 (0.518-0.531) | 0.708 |
| Left Axis Deviation | 0.940 | 0.649 | 0.975 | 0.959 | 0.753 | 0.856 (0.853-0.859) | 0.511 (0.504-0.518) | 0.697 |
| Acute MI* | 0.985 | 0.255 | 0.998 | 0.986 | 0.711 | 0.849 (0.838-0.86) | 0.183 (0.171-0.194) | 0.375 |
| Lead Reversal | 0.997 | 0.325 | 1.000 | 0.997 | 0.718 | 0.858 (0.836-0.879) | 0.234 (0.206-0.261) | 0.447 |
| 1<sup>st</sup> degree AVb | 0.964 | 0.743 | 0.983 | 0.978 | 0.787 | 0.883 (0.88-0.886) | 0.601 (0.594-0.608) | 0.764 |
| 2<sup>nd</sup> degree AVb | 0.998 | 0.379 | 0.999 | 0.999 | 0.643 | 0.821 (0.796-0.846) | 0.244 (0.207-0.283) | 0.477 |
| 3<sup>rd</sup> degree AVb | 0.998 | 0.260 | 1.000 | 0.998 | 0.693 | 0.846 (0.816-0.875) | 0.181 (0.149-0.21) | 0.378 |
| WPW | 0.999 | 0.444 | 1.000 | 1.000 | 0.624 | 0.812 (0.771-0.853) | 0.277 (0.218-0.353) | 0.519 |
| Repol Abnormality | 0.930 | 0.515 | 0.973 | 0.951 | 0.661 | 0.806 (0.802-0.809) | 0.365 (0.359-0.37) | 0.579 |

Furthermore, ECG-GPT performed comparably to two previously published multilabel CNN models^14,42^ across common labels (**Table S5**). There was no significant difference in the performance for the labels of interest when comparing ECGs recorded in the inpatient, emergency, and outpatient setting, with median AUROCs of 0.89 (IQR 0.85-0.92), 0.90 (IQR 0.87-0.94), and 0.89 (IQR 0.83-0.94), respectively (p=0.696).

### External Validation – Mount Sinai Health System

We used a total of 1,434,455 ECG images generated from raw 12-lead ECG signal data and 10,052 prospectively acquired native ECG images from MSHS for validation. In assessing semantic similarity for the set of generated ECG images, embeddings had a median pairwise similarity of 0.86 (IQR 0.79-0.96), significantly greater than the median baseline similarity of 0.74 among 2 random statements (IQR 0.69-0.80, p<0.001). For the set of native ECG images, embeddings had a median pairwise and baseline similarity of 0.80 (IQR 0.72-0.86) and 0.73 (IQR 0.68-0.78), respectively. This separation persisted across the 26 subsets corresponding to each extracted rhythm and conduction disorder (**Table 3** and **Table 4**). For key rhythm disorders – AF, ST, SB, PACs, and PVCs – pairwise and baseline similarity ranged from 0.86-0.89 and 0.78-0.81, respectively, for the generated ECG images and 0.78-0.87 and 0.75-0.80, respectively, for the native ECG images. For key conduction abnormalities – LBBB, RBBB, 1dAVb, LAFB, and LFPB – pairwise and baseline similarity ranged from 0.88-0.95 and 0.79-0.85, respectively, for the generated ECG images and 0.84-0.85 and 0.77-0.81, respectively, for the native ECG images.

**Table 3.** Pairwise and baseline similarity between reference and model-generated statements for plotted ECG signals in the Mount Sinai Health System. Abbreviations: AF, atrial fibrillation; ST, sinus tachycardia; SB, sinus bradycardia; LBBB, left bundle branch block; RBBB, right bundle branch block; LAFB, left anterior fascicular block; LPFB, left posterior fascicular block; SVT, supraventricular tachycardia; PAC, premature atrial complexes; PVC, premature ventricular complexes; LAE, left atrial enlargement; RAE, right atrial enlargement; LVH, left ventricular hypertrophy; RVH, right ventricular hypertrophy; MI, myocardial infarction; AVb, atrioventricular block; WPW, Wolff-Parkinson-White syndrome; Repol, repolarization. *Acute MI includes ST Elevation MI (STEMI).

| Labels | Pairwise Similarity | Baseline Similarity | P-Value |
| --- | --- | --- | --- |
| Overall | 0.861 (0.786-0.961) | 0.740 (0.687-0.795) | <0.001 |
| Sinus Rhythm | 0.853 (0.777-0.964) | 0.751 (0.696-0.806) | <0.001 |
| AF | 0.880 (0.811-0.946) | 0.807 (0.764-0.852) | <0.001 |
| Atrial Flutter | 0.871 (0.807-0.933) | 0.811 (0.762-0.854) | <0.001 |
| ST | 0.857 (0.790-0.938) | 0.780 (0.733-0.830) | <0.001 |
| SB | 0.904 (0.814-1.000) | 0.794 (0.738-0.845) | <0.001 |
| Sinus Arrhythmia | 0.918 (0.794-1.000) | 0.829 (0.744-0.890) | <0.001 |
| LBBB | 0.945 (0.854-1.000) | 0.853 (0.798-0.897) | <0.001 |
| RBBB | 0.897 (0.839-0.952) | 0.812 (0.770-0.853) | <0.001 |
| LAFB | 0.900 (0.838-0.948) | 0.808 (0.766-0.848) | <0.001 |
| LPFB | 0.877 (0.811-0.932) | 0.793 (0.745-0.839) | <0.001 |
| SVT | 0.831 (0.763-0.900) | 0.783 (0.723-0.835) | <0.001 |
| PAC | 0.858 (0.799-0.922) | 0.785 (0.740-0.831) | <0.001 |
| PVC | 0.893 (0.816-0.958) | 0.807 (0.758-0.855) | <0.001 |
| LAE | 0.916 (0.816-0.984) | 0.815 (0.752-0.866) | <0.001 |
| RAE | 0.884 (0.808-0.963) | 0.795 (0.743-0.841) | <0.001 |
| LVH | 0.922 (0.819-0.978) | 0.825 (0.770-0.877) | <0.001 |
| RVH | 0.832 (0.774-0.904) | 0.778 (0.732-0.827) | <0.001 |
| Low Voltage | 0.840 (0.761-0.927) | 0.764 (0.707-0.824) | <0.001 |
| Left Axis Deviation | 0.891 (0.810-0.957) | 0.785 (0.737-0.829) | <0.001 |
| Acute MI | 0.796 (0.730-0.926) | 0.760 (0.706-0.880) | <0.001 |
| Lead Reversal | 0.847 (0.736-0.902) | 0.789 (0.686-0.866) | <0.001 |
| 1st degree AVb | 0.907 (0.817-0.968) | 0.819 (0.760-0.867) | <0.001 |
| 2nd degree AVb | 0.871 (0.797-0.941) | 0.812 (0.751-0.870) | <0.001 |
| 3rd degree AVb | 0.817 (0.743-0.880) | 0.767 (0.712-0.820) | <0.001 |
| WPW | 0.748 (0.671-0.942) | 0.707 (0.660-0.808) | <0.001 |
| Repol Abnormality | 0.875 (0.810-0.954) | 0.793 (0.719-0.856) | <0.001 |

**Table 4.**
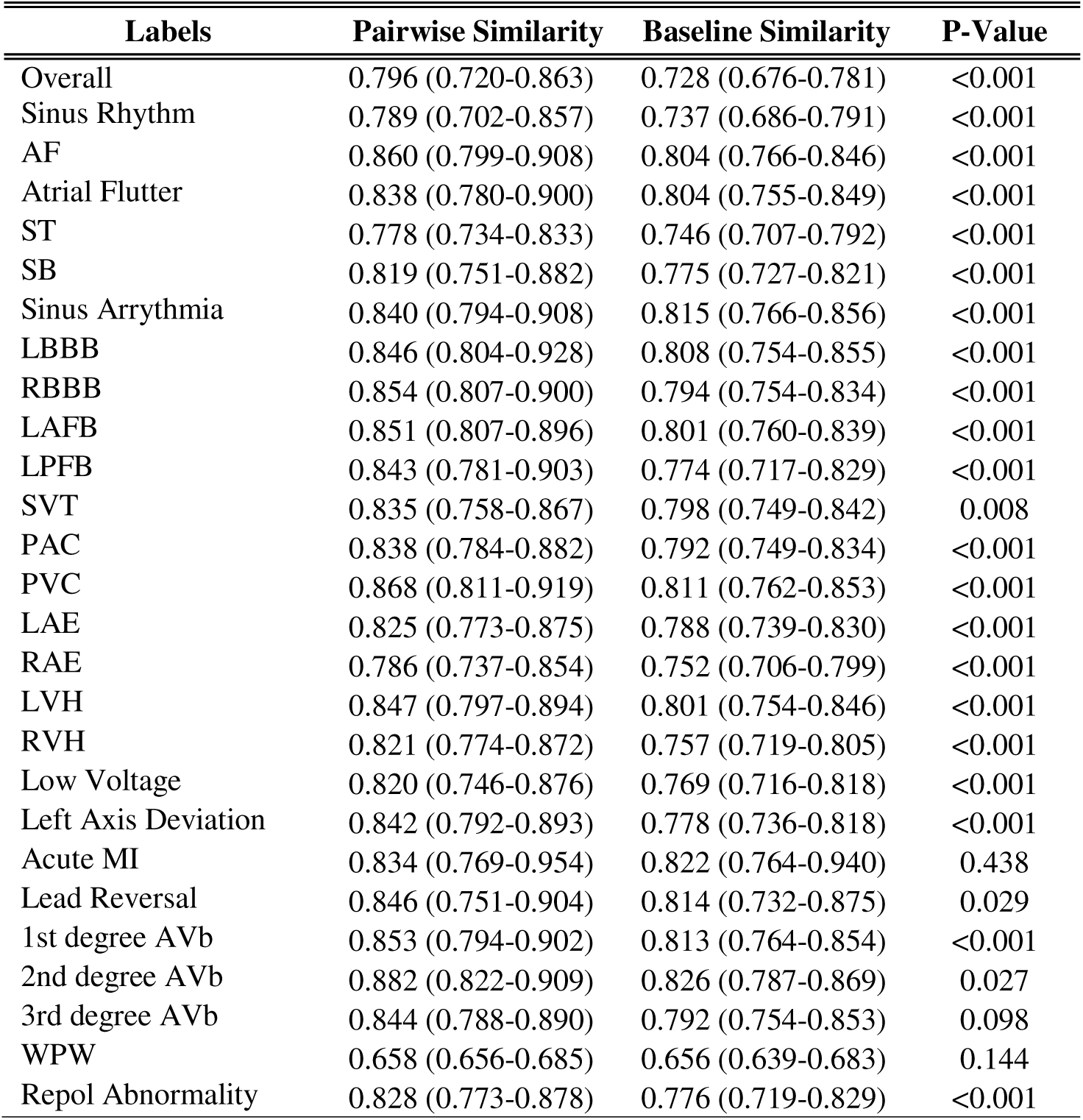
Pairwise and baseline similarity between reference and model-generated statements for prospectively acquired PDFs of ECGs in the Mount Sinai Health System. Abbreviations: AF, atrial fibrillation; ST, sinus tachycardia; SB, sinus bradycardia; LBBB, left bundle branch block; RBBB, right bundle branch block; LAFB, left anterior fascicular block; LPFB, left posterior fascicular block; SVT, supraventricular tachycardia; PAC, premature atrial complexes; PVC, premature ventricular complexes; LAE, left atrial enlargement; RAE, right atrial enlargement; LVH, left ventricular hypertrophy; RVH, right ventricular hypertrophy; MI, myocardial infarction; AVb, atrioventricular block; WPW, Wolff-Parkinson-White syndrome; Repol, repolarization. *Acute MI includes ST Elevation MI (STEMI).

The model performed well in clinical assessment across the 26 extracted labels. For AF, ST, SB, PACs, and PVCs, AUROCs and AUPRCs ranged from 0.74-0.94 and 0.40-0.83, respectively, for the generated ECG images and 0.67-0.91 and 0.30-0.76, respectively, for the native ECG images. For LBBB, RBBB, 1dAVb, LAFB, and LFPB, AUROCs and AUPRCs ranged from 0.87-0.97 and 0.14-0.83, respectively, for the generated ECG images and 0.71-0.97 and 0.05-0.63, respectively, for the native ECG images. Model performance in the native image MSHS validation dataset across all 26 conditions is reported in **Table 5** and **Table 6**.

**Table 5.** Clinical assessment of model-generated ECG interpretation statements on plotted ECG signals in the Mount Sinai Health System. Abbreviations: PPV, Positive Predictive Value; NPV, Negative Predictive Value; AUROC, area under the receiver operator characteristic; AUPRC, area under precision-recall curve; AF, atrial fibrillation; ST, sinus tachycardia; SB, sinus bradycardia; LBBB, left bundle branch block; RBBB, right bundle branch block; LAFB, left anterior fascicular block; LPFB, left posterior fascicular block; SVT, supraventricular tachycardia; PAC, premature atrial complexes; PVC, premature ventricular complexes; LAE, left atrial enlargement; RAE, right atrial enlargement; LVH, left ventricular hypertrophy; RVH, right ventricular hypertrophy; MI, myocardial infarction; AVb, atrioventricular block; WPW, Wolff-Parkinson-White syndrome; Repol, repolarization. *Acute MI includes ST Elevation MI (STEMI).

| Labels | Accuracy | PPV | NPV | Specificity | Sensitivity | AUROC | AUPRC | F1 |
| --- | --- | --- | --- | --- | --- | --- | --- | --- |
| Sinus Rhythm | 0.931 | 0.942 | 0.908 | 0.883 | 0.955 | 0.919 (0.919-0.92) | 0.93 (0.929-0.93) | 0.949 |
| AF | 0.978 | 0.89 | 0.984 | 0.992 | 0.793 | 0.893 (0.891-0.894) | 0.721 (0.718-0.724) | 0.839 |
| Atrial Flutter | 0.975 | 0.372 | 0.997 | 0.978 | 0.838 | 0.908 (0.905-0.91) | 0.314 (0.31-0.318) | 0.515 |
| ST | 0.979 | 0.91 | 0.988 | 0.989 | 0.898 | 0.944 (0.943-0.944) | 0.828 (0.827-0.83) | 0.904 |
| SB | 0.965 | 0.948 | 0.967 | 0.995 | 0.734 | 0.865 (0.863-0.866) | 0.726 (0.724-0.728) | 0.828 |
| Sinus Arrhythmia | 0.941 | 0.412 | 0.987 | 0.951 | 0.728 | 0.84 (0.838-0.841) | 0.312 (0.309-0.315) | 0.526 |
| LBBB | 0.99 | 0.746 | 0.998 | 0.992 | 0.914 | 0.953 (0.952-0.954) | 0.684 (0.679-0.688) | 0.821 |
| RBBB | 0.987 | 0.876 | 0.996 | 0.99 | 0.94 | 0.965 (0.965-0.966) | 0.827 (0.825-0.829) | 0.907 |
| LAFB | 0.969 | 0.558 | 0.991 | 0.977 | 0.776 | 0.877 (0.875-0.878) | 0.441 (0.437-0.445) | 0.649 |
| LPFB | 0.985 | 0.179 | 0.999 | 0.986 | 0.753 | 0.869 (0.864-0.875) | 0.136 (0.131-0.141) | 0.289 |
| SVT | 0.993 | 0.279 | 0.999 | 0.995 | 0.644 | 0.819 (0.812-0.826) | 0.181 (0.172-0.188) | 0.389 |
| PAC | 0.961 | 0.763 | 0.968 | 0.991 | 0.479 | 0.735 (0.733-0.737) | 0.396 (0.392-0.399) | 0.589 |
| PVC | 0.972 | 0.749 | 0.988 | 0.981 | 0.826 | 0.904 (0.902-0.905) | 0.629 (0.626-0.633) | 0.785 |
| LAE | 0.923 | 0.564 | 0.986 | 0.927 | 0.877 | 0.902 (0.901-0.903) | 0.506 (0.504-0.508) | 0.686 |
| RAE | 0.985 | 0.284 | 0.999 | 0.986 | 0.839 | 0.913 (0.909-0.916) | 0.239 (0.234-0.245) | 0.424 |
| LVH | 0.951 | 0.749 | 0.988 | 0.955 | 0.922 | 0.938 (0.938-0.939) | 0.701 (0.699-0.703) | 0.827 |
| RVH | 0.937 | 0.084 | 0.996 | 0.94 | 0.607 | 0.774 (0.769-0.778) | 0.054 (0.053-0.056) | 0.147 |
| Low Voltage | 0.957 | 0.592 | 0.988 | 0.966 | 0.813 | 0.89 (0.888-0.891) | 0.493 (0.49-0.495) | 0.686 |
| Left Axis Deviation | 0.934 | 0.639 | 0.967 | 0.96 | 0.687 | 0.824 (0.822-0.825) | 0.469 (0.466-0.471) | 0.663 |
| Acute MI* | 0.974 | 0.22 | 0.997 | 0.977 | 0.672 | 0.825 (0.821-0.829) | 0.151 (0.146-0.154) | 0.331 |
| Lead Reversal | 0.993 | 0.175 | 0.999 | 0.994 | 0.512 | 0.753 (0.745-0.761) | 0.091 (0.085-0.097) | 0.261 |
| 1<sup>st</sup> degree AVb | 0.97 | 0.699 | 0.99 | 0.977 | 0.845 | 0.911 (0.91-0.912) | 0.599 (0.596-0.603) | 0.765 |
| 2<sup>nd</sup> degree AVb | 0.998 | 0.421 | 0.999 | 0.998 | 0.563 | 0.781 (0.772-0.79) | 0.238 (0.22-0.253) | 0.482 |
| 3<sup>rd</sup> degree AVb | 0.998 | 0.277 | 1 | 0.999 | 0.655 | 0.827 (0.813-0.84) | 0.182 (0.167-0.195) | 0.389 |
| WPW | 0.999 | 0.533 | 1 | 1 | 0.608 | 0.804 (0.789-0.819) | 0.325 (0.302-0.354) | 0.568 |
| Repol Abnormality | 0.927 | 0.319 | 0.985 | 0.938 | 0.667 | 0.803 (0.801-0.805) | 0.227 (0.225-0.229) | 0.432 |

**Table 6.** Clinical assessment of model-generated ECG interpretation statements on prospectively acquired PDFs of ECGs in the Mount Sinai Health System. Abbreviations: PPV, Positive Predictive Value; NPV, Negative Predictive Value; AUROC, area under the receiver operator characteristic; AUPRC, area under precision-recall curve; AF, atrial fibrillation; ST, sinus tachycardia; SB, sinus bradycardia; LBBB, left bundle branch block; RBBB, right bundle branch block; LAFB, left anterior fascicular block; LPFB, left posterior fascicular block; SVT, supraventricular tachycardia; PAC, premature atrial complexes; PVC, premature ventricular complexes; LAE, left atrial enlargement; RAE, right atrial enlargement; LVH, left ventricular hypertrophy; RVH, right ventricular hypertrophy; MI, myocardial infarction; AVb, atrioventricular block; WPW, Wolff-Parkinson-White syndrome; Repol, repolarization. *Acute MI includes ST Elevation MI (STEMI)

| Labels | Accuracy | PPV | NPV | Specificity | Sensitivity | AUROC | AUPRC | F1 |
| --- | --- | --- | --- | --- | --- | --- | --- | --- |
| Sinus Rhythm | 0.92 | 0.946 | 0.879 | 0.909 | 0.927 | 0.918 (0.912-0.924) | 0.923 (0.917-0.929) | 0.936 |
| AF | 0.96 | 0.693 | 0.986 | 0.971 | 0.821 | 0.896 (0.882-0.91) | 0.582 (0.55-0.615) | 0.752 |
| Atrial Flutter | 0.958 | 0.207 | 0.996 | 0.961 | 0.735 | 0.848 (0.811-0.886) | 0.156 (0.126-0.189) | 0.323 |
| ST | 0.959 | 0.797 | 0.978 | 0.977 | 0.804 | 0.89 (0.878-0.903) | 0.661 (0.634-0.685) | 0.8 |
| SB | 0.97 | 0.895 | 0.978 | 0.988 | 0.822 | 0.905 (0.894-0.916) | 0.755 (0.732-0.781) | 0.857 |
| Sinus Arrhythmia | 0.909 | 0.325 | 0.993 | 0.911 | 0.865 | 0.888 (0.872-0.904) | 0.288 (0.265-0.315) | 0.473 |
| LBBB | 0.979 | 0.559 | 0.997 | 0.981 | 0.898 | 0.939 (0.921-0.958) | 0.504 (0.45-0.555) | 0.689 |
| RBBB | 0.961 | 0.645 | 0.998 | 0.96 | 0.973 | 0.966 (0.96-0.973) | 0.629 (0.597-0.655) | 0.776 |
| LAFB | 0.956 | 0.415 | 0.979 | 0.976 | 0.453 | 0.714 (0.689-0.74) | 0.208 (0.176-0.246) | 0.433 |
| LPFB | 0.977 | 0.084 | 0.999 | 0.978 | 0.625 | 0.802 (0.716-0.887) | 0.054 (0.023-0.086) | 0.148 |
| SVT | 0.985 | 0.206 | 0.999 | 0.986 | 0.841 | 0.913 (0.859-0.968) | 0.174 (0.117-0.227) | 0.33 |
| PAC | 0.95 | 0.738 | 0.957 | 0.992 | 0.347 | 0.669 (0.651-0.688) | 0.298 (0.264-0.336) | 0.472 |
| PVC | 0.963 | 0.694 | 0.986 | 0.975 | 0.801 | 0.888 (0.873-0.903) | 0.57 (0.539-0.605) | 0.744 |
| LAE | 0.9 | 0.436 | 0.967 | 0.922 | 0.654 | 0.788 (0.772-0.804) | 0.314 (0.288-0.339) | 0.523 |
| RAE | 0.988 | 0.235 | 0.996 | 0.992 | 0.369 | 0.681 (0.622-0.74) | 0.091 (0.042-0.145) | 0.287 |
| LVH | 0.813 | 0.377 | 0.927 | 0.851 | 0.572 | 0.712 (0.698-0.725) | 0.274 (0.256-0.292) | 0.455 |
| RVH | 0.925 | 0.023 | 0.997 | 0.928 | 0.378 | 0.653 (0.581-0.724) | 0.011 (0.006-0.019) | 0.043 |
| Low Voltage | 0.948 | 0.518 | 0.978 | 0.967 | 0.617 | 0.792 (0.771-0.812) | 0.34 (0.304-0.379) | 0.563 |
| Left Axis Deviation | 0.917 | 0.605 | 0.949 | 0.959 | 0.551 | 0.755 (0.739-0.77) | 0.379 (0.351-0.408) | 0.577 |
| Acute MI* | 0.89 | 0.04 | 0.998 | 0.891 | 0.75 | 0.82 (0.765-0.876) | 0.031 (0.021-0.041) | 0.075 |
| Lead Reversal | 0.96 | 0.052 | 0.998 | 0.962 | 0.525 | 0.743 (0.665-0.822) | 0.029 (0.015-0.049) | 0.095 |
| 1<sup>st</sup> degree AVb | 0.942 | 0.55 | 0.988 | 0.949 | 0.843 | 0.896 (0.882-0.91) | 0.475 (0.441-0.508) | 0.666 |
| 2<sup>nd</sup> degree AVb | 0.996 | 0.225 | 0.999 | 0.997 | 0.45 | 0.723 (0.612-0.835) | 0.102 (0.032-0.196) | 0.3 |
| 3<sup>rd</sup> degree AVb | 0.996 | 0.216 | 0.999 | 0.997 | 0.471 | 0.734 (0.612-0.856) | 0.103 (0.028-0.211) | 0.296 |
| WPW | 0.999 | 0.333 | 0.999 | 1 | 0.143 | 0.571 (0.431-0.711) | 0.048 (0-0.334) | 0.2 |
| Repol Abnormality | 0.908 | 0.198 | 0.98 | 0.924 | 0.497 | 0.711 (0.685-0.736) | 0.117 (0.101-0.14) | 0.283 |

### External Validation – Houston Methodist Hospital

We also deployed ECG-GPT in a set of 62,876 ECGs in their native image format from HMH. Across conditions, embeddings had a median pairwise similarity of 0.81 (IQR 0.75-0.88), significantly greater than the median baseline similarity of 0.77 among 2 random statements (IQR 0.73-0.82, p<0.001). This separation persisted across the majority of the 26 subsets corresponding to each extracted rhythm and conduction disorder (**Table 7**). In clinical assessment, AUROCs and AUPRCs, respectively, for AF, ST, SB, PACs, and PVCs ranged from 0.65-0.96 and 0.13-0.68. For LBBB, RBBB, 1dAVb, LAFB, and LFPB, AUROCs and AUPRCs ranged from 0.74-0.97 and 0.04-0.77, respectively. Model performance across all 26 conditions is reported in **Table 8**.

**Table 7.** Pairwise and baseline similarity between reference and model-generated statements in the Houston Methodist Hospital. Abbreviations: AF, atrial fibrillation; ST, sinus tachycardia; SB, sinus bradycardia; LBBB, left bundle branch block; RBBB, right bundle branch block; LAFB, left anterior fascicular block; LPFB, left posterior fascicular block; SVT, supraventricular tachycardia; PAC, premature atrial complexes; PVC, premature ventricular complexes; LAE, left atrial enlargement; RAE, right atrial enlargement; LVH, left ventricular hypertrophy; RVH, right ventricular hypertrophy; MI, myocardial infarction; AVb, atrioventricular block; WPW, Wolff-Parkinson-White syndrome; Repol, repolarization. *Acute MI includes ST Elevation MI (STEMI).

| Labels | Pairwise Similarity | Baseline Similarity | P-Value |
| --- | --- | --- | --- |
| Overall | 0.814 (0.753-0.882) | 0.772 (0.733-0.817) | <0.001 |
| Sinus Rhythm | 0.797 (0.747-0.873) | 0.772 (0.735-0.818) | <0.001 |
| AF | 0.884 (0.840-0.922) | 0.855 (0.817-0.889) | <0.001 |
| Atrial Flutter | 0.878 (0.832-0.919) | 0.860 (0.814-0.896) | <0.001 |
| ST | 0.856 (0.808-0.902) | 0.826 (0.790-0.862) | <0.001 |
| SB | 0.828 (0.774-0.888) | 0.800 (0.758-0.842) | <0.001 |
| Sinus Arrhythmia | 0.892 (0.854-0.939) | 0.866 (0.832-0.899) | <0.001 |
| LBBB | 0.902 (0.865-0.982) | 0.865 (0.824-0.910) | <0.001 |
| RBBB | 0.903 (0.865-0.948) | 0.853 (0.815-0.886) | <0.001 |
| LAFB | 0.895 (0.862-0.930) | 0.851 (0.813-0.882) | <0.001 |
| LPFB | 0.867 (0.830-0.903) | 0.823 (0.784-0.860) | <0.001 |
| SVT | 0.880 (0.828-0.921) | 0.851 (0.813-0.886) | <0.001 |
| PAC | 0.871 (0.825-0.915) | 0.836 (0.796-0.873) | <0.001 |
| PVC | 0.909 (0.857-0.952) | 0.862 (0.820-0.898) | <0.001 |
| LAE | 0.879 (0.838-0.923) | 0.859 (0.828-0.889) | <0.001 |
| RAE | 0.869 (0.831-0.921) | 0.854 (0.823-0.885) | <0.001 |
| LVH | 0.913 (0.885-0.943) | 0.880 (0.850-0.906) | <0.001 |
| RVH | 0.897 (0.854-0.934) | 0.854 (0.817-0.890) | <0.001 |
| Low Voltage | 0.838 (0.797-0.885) | 0.814 (0.782-0.850) | <0.001 |
| Left Axis Deviation | 0.890 (0.848-0.926) | 0.841 (0.808-0.872) | <0.001 |
| Acute MI | 0.880 (0.840-0.909) | 0.865 (0.832-0.895) | 0.007 |
| Lead Reversal | 0.909 (0.863-0.935) | 0.880 (0.827-0.910) | <0.001 |
| 1st degree AVb | 0.895 (0.839-0.939) | 0.864 (0.817-0.901) | <0.001 |
| 2nd degree AVb | 0.902 (0.850-0.953) | 0.890 (0.852-0.924) | 0.041 |
| 3rd degree AVb | 0.829 (0.779-0.883) | 0.823 (0.780-0.872) | 0.547 |
| WPW | 0.777 (0.755-0.871) | 0.786 (0.755-0.866) | 0.94 |
| Repol Abnormality | 0.878 (0.828-0.931) | 0.833 (0.789-0.878) | <0.001 |

**Table 8.** Clinical assessment of model-generated ECG interpretation statements on PDFs of ECGs in the Houston Methodist Hospital. Abbreviations: PPV, Positive Predictive Value; NPV, Negative Predictive Value; AUROC, area under the receiver operator characteristic; AUPRC, area under precision-recall curve; AF, atrial fibrillation; ST, sinus tachycardia; SB, sinus bradycardia; LBBB, left bundle branch block; RBBB, right bundle branch block; LAFB, left anterior fascicular block; LPFB, left posterior fascicular block; SVT, supraventricular tachycardia; PAC, premature atrial complexes; PVC, premature ventricular complexes; LAE, left atrial enlargement; RAE, right atrial enlargement; LVH, left ventricular hypertrophy; RVH, right ventricular hypertrophy; MI, myocardial infarction; AVb, atrioventricular block; WPW, Wolff-Parkinson-White syndrome; Repol, repolarization. *Acute MI includes ST Elevation MI (STEMI).

| Labels | Accuracy | PPV | NPV | Specificity | Sensitivity | AUROC | AUPRC | F1 |
| --- | --- | --- | --- | --- | --- | --- | --- | --- |
| Sinus Rhythm | 0.888 | 0.89 | 0.881 | 0.701 | 0.962 | 0.832 (0.828-0.835) | 0.883 (0.881-0.886) | 0.925 |
| AF | 0.989 | 0.801 | 0.995 | 0.994 | 0.837 | 0.916 (0.907-0.924) | 0.675 (0.652-0.698) | 0.819 |
| Atrial Flutter | 0.989 | 0.312 | 0.999 | 0.990 | 0.839 | 0.914 (0.895-0.934) | 0.263 (0.233-0.298) | 0.455 |
| ST | 0.976 | 0.596 | 0.998 | 0.978 | 0.937 | 0.957 (0.952-0.962) | 0.561 (0.542-0.579) | 0.729 |
| SB | 0.923 | 0.977 | 0.917 | 0.997 | 0.589 | 0.793 (0.788-0.797) | 0.65 (0.642-0.656) | 0.735 |
| Sinus Arrhythmia | 0.956 | 0.516 | 0.995 | 0.958 | 0.905 | 0.932 (0.926-0.937) | 0.471 (0.457-0.486) | 0.657 |
| LBBB | 0.996 | 0.767 | 0.999 | 0.997 | 0.905 | 0.951 (0.94-0.962) | 0.696 (0.67-0.73) | 0.831 |
| RBBB | 0.989 | 0.809 | 0.998 | 0.991 | 0.944 | 0.967 (0.963-0.972) | 0.765 (0.752-0.78) | 0.871 |
| LAFB | 0.975 | 0.451 | 0.993 | 0.981 | 0.698 | 0.84 (0.827-0.852) | 0.321 (0.298-0.345) | 0.548 |
| LPFB | 0.982 | 0.082 | 0.998 | 0.984 | 0.497 | 0.74 (0.704-0.777) | 0.042 (0.03-0.056) | 0.141 |
| SVT | 0.998 | 0.497 | 0.999 | 0.998 | 0.722 | 0.86 (0.822-0.898) | 0.36 (0.3-0.431) | 0.589 |
| PAC | 0.976 | 0.798 | 0.978 | 0.997 | 0.308 | 0.653 (0.642-0.663) | 0.267 (0.249-0.289) | 0.444 |
| PVC | 0.791 | 0.139 | 0.995 | 0.787 | 0.892 | 0.84 (0.833-0.846) | 0.128 (0.123-0.133) | 0.241 |
| LAE | 0.880 | 0.258 | 0.994 | 0.879 | 0.896 | 0.887 (0.882-0.893) | 0.235 (0.227-0.244) | 0.400 |
| RAE | 0.961 | 0.141 | 0.999 | 0.962 | 0.904 | 0.933 (0.919-0.947) | 0.128 (0.113-0.143) | 0.244 |
| LVH | 0.852 | 0.265 | 0.996 | 0.847 | 0.943 | 0.895 (0.891-0.899) | 0.253 (0.246-0.261) | 0.413 |
| RVH | 0.940 | 0.028 | 0.999 | 0.941 | 0.644 | 0.793 (0.756-0.83) | 0.019 (0.014-0.024) | 0.053 |
| Low Voltage | 0.953 | 0.664 | 0.967 | 0.983 | 0.498 | 0.74 (0.733-0.748) | 0.362 (0.347-0.379) | 0.569 |
| Left Axis Deviation | 0.950 | 0.543 | 0.978 | 0.969 | 0.623 | 0.796 (0.788-0.804) | 0.359 (0.345-0.372) | 0.580 |
| Acute MI* | 0.943 | 0.026 | 1.000 | 0.943 | 0.835 | 0.889 (0.855-0.923) | 0.022 (0.016-0.027) | 0.051 |
| Lead Reversal | 0.767 | 0.005 | 0.999 | 0.768 | 0.693 | 0.73 (0.685-0.776) | 0.004 (0.003-0.005) | 0.009 |
| 1<sup>st</sup> degree AVb | 0.980 | 0.809 | 0.988 | 0.991 | 0.755 | 0.873 (0.866-0.881) | 0.622 (0.606-0.642) | 0.781 |
| 2<sup>nd</sup> degree AVb | 0.999 | 0.484 | 1.000 | 0.999 | 0.608 | 0.804 (0.736-0.871) | 0.295 (0.18-0.413) | 0.539 |
| 3<sup>rd</sup> degree AVb | 1.000 | 0.481 | 1.000 | 1.000 | 0.464 | 0.732 (0.638-0.826) | 0.224 (0.089-0.388) | 0.473 |
| WPW | 0.998 | 0.129 | 1.000 | 0.998 | 0.652 | 0.825 (0.726-0.925) | 0.084 (0.035-0.147) | 0.216 |
| Repol Abnormality | 0.931 | 0.115 | 0.994 | 0.935 | 0.6 | 0.768 (0.751-0.784) | 0.074 (0.066-0.084) | 0.193 |

### External Validation – Lake Regional Hospital

Next, we used a real-world dataset of 64 ECG images collected at LRH in Osage Beach, MO. In these ECGs, the model had AUROCs of 0.95, 0.95, and 0.75 for AF, ST, and SB, respectively. For LBBB, RBBB, and 1dAVb, the model reported AUROCs of 0.82, 1.00, and 0.87, respectively.

### External Validation – Open-source datasets

The model’s diagnostic performance in each of the four open-source datasets used for validation is noted in **Table 9**. First, in 2,228,236 ECGs from the previously described CODE15 dataset collected by the Telehealth Network of Minas Gerais (TNMG), Brazil, between 2010 and 2017.^12,23^ Here, AUROCs for rhythm disorders were 0.94, 0.92, and 0.92, and AUPRCs were 0.56, 0.54, and 0.11, for AF, ST, and SB, respectively. The model performed similarly for conduction abnormalities, with AUROCs of 0.90, 0.96, and 0.89, and AUPRCs of 0.62, 0.67, and 0.29 for LBBB, RBBB, and 1dAVb, respectively. When deployed to a smaller, cardiologist-validated dataset collected by TNMG in Brazil between April and September 2018, consisting of 827 ECGs manually annotated by two cardiologists with disputes resolved by a third,^15^ AUROCs were higher across nearly all diagnostic labels. The model reported AUROCs of 1.00 (rounded from 0.998), 0.98, and 0.94 for AF, ST, and SB, respectively. For LBBB, RBBB, and 1dAVb, the model reported AUROCs of 0.94, 0.97, and 0.85, respectively.

**Table 9.** Clinical assessment of model-generated ECG interpretation statements on external validation sets. Abbreviations: ECG, electrocardiogram; PPV, Positive Predictive Value; NPV, Negative Predictive Value; Spec, specificity; Sens, sensitivity; AUROC, area under the receiver operator characteristic; AUPRC, area under precision-recall curve. AF, atrial fibrillation; ST, sinus tachycardia; SB, sinus bradycardia; LBBB, left bundle branch block; RBBB, right bundle branch block; AVb, atrioventricular block.

|  | Labels | Accuracy | PPV | NPV | Specificity | Sensitivity | AUROC | AUPRC | F1 |
| --- | --- | --- | --- | --- | --- | --- | --- | --- | --- |
| Cardiologist Validated CODE15 | AF | 0.996 | 0.765 | 1.000 | 0.995 | 1.000 | 0.998 (0.996-1.00) | 0.765 (0.550-0.947) | 0.867 |
|  | ST | 0.993 | 0.884 | 0.999 | 0.994 | 0.974 | 0.984 (0.959-1) | 0.862 (0.749-0.973) | 0.927 |
|  | SB | 0.890 | 0.139 | 1.000 | 0.888 | 1.000 | 0.944 (0.934-0.954) | 0.139 (0.077-0.202) | 0.244 |
|  | LBBB | 0.996 | 1.000 | 0.995 | 1.000 | 0.871 | 0.935 (0.876-0.995) | 0.875 (0.759-0.971) | 0.931 |
|  | RBBB | 0.990 | 0.829 | 0.998 | 0.992 | 0.944 | 0.968 (0.93-1) | 0.785 (0.639-0.898) | 0.883 |
|  | AVb | 0.976 | 0.629 | 0.990 | 0.985 | 0.710 | 0.847 (0.766-0.929) | 0.456 (0.293-0.603) | 0.667 |
| CODE15 | AF | 0.988 | 0.618 | 0.998 | 0.990 | 0.897 | 0.943 (0.942-0.945) | 0.556 (0.551-0.559) | 0.731 |
|  | ST | 0.986 | 0.625 | 0.997 | 0.989 | 0.855 | 0.922 (0.92-0.923) | 0.538 (0.534-0.543) | 0.722 |
|  | SB | 0.874 | 0.109 | 0.999 | 0.872 | 0.972 | 0.922 (0.921-0.923) | 0.107 (0.106-0.108) | 0.197 |
|  | LBBB | 0.993 | 0.766 | 0.997 | 0.996 | 0.810 | 0.903 (0.901-0.905) | 0.624 (0.619-0.629) | 0.788 |
|  | RBBB | 0.988 | 0.724 | 0.998 | 0.990 | 0.923 | 0.956 (0.955-0.957) | 0.670 (0.667-0.673) | 0.811 |
|  | AVb | 0.974 | 0.353 | 0.997 | 0.977 | 0.811 | 0.894 (0.892-0.896) | 0.289 (0.286-0.293) | 0.492 |
| UK Biobank | AF | 0.997 | 0.939 | 0.998 | 0.999 | 0.876 | 0.937 (0.925-0.95) | 0.824 (0.799-0.855) | 0.906 |
|  | ST | 0.999 | 0.862 | 1.000 | 1.000 | 0.815 | 0.907 (0.868-0.947) | 0.703 (0.599-0.798) | 0.838 |
|  | SB | 0.924 | 0.973 | 0.894 | 0.981 | 0.853 | 0.917 (0.914-0.919) | 0.894 (0.890-0.898) | 0.909 |
|  | LBBB | 0.999 | 0.871 | 1.000 | 0.999 | 0.982 | 0.991 (0.984-0.997) | 0.855 (0.822-0.884) | 0.923 |
|  | RBBB | 0.996 | 0.832 | 1.000 | 0.996 | 0.979 | 0.987 (0.983-0.992) | 0.814 (0.791-0.840) | 0.899 |
|  | AVb | 0.969 | 0.663 | 0.995 | 0.973 | 0.911 | 0.942 (0.936-0.947) | 0.609 (0.592-0.627) | 0.767 |
| PTB-XL | AF | 0.978 | 0.787 | 0.995 | 0.981 | 0.931 | 0.956 (0.949-0.962) | 0.738 (0.716-0.758) | 0.853 |
|  | ST | 0.990 | 0.836 | 0.996 | 0.993 | 0.907 | 0.950 (0.940-0.960) | 0.762 (0.732-0.787) | 0.870 |
|  | SB | 0.876 | 0.184 | 0.998 | 0.874 | 0.942 | 0.908 (0.899-0.917) | 0.175 (0.161-0.187) | 0.308 |
|  | LBBB | 0.995 | 0.857 | 0.999 | 0.996 | 0.944 | 0.970 (0.960-0.980) | 0.811 (0.783-0.846) | 0.899 |
|  | RBBB | 0.989 | 0.691 | 0.999 | 0.989 | 0.980 | 0.984 (0.978-0.99) | 0.678 (0.645-0.713) | 0.810 |
|  | AVb | 0.954 | 0.419 | 0.989 | 0.963 | 0.711 | 0.837 (0.821-0.853) | 0.309 (0.284-0.335) | 0.528 |
| LRH | AFIB | 0.969 | 0.923 | 0.980 | 0.980 | 0.923 | 0.952 (0.874-1.00) | 0.868 (0.665-1.00) | 0.923 |
|  | ST | 0.984 | 1.000 | 0.982 | 1.000 | 0.900 | 0.950 (0.852-1.00) | 0.916 (0.714-1.00) | 0.947 |
|  | SB | 0.922 | 1.000 | 0.915 | 1.000 | 0.500 | 0.750 (0.587-0.913) | 0.578 (0.245-0.873) | 0.667 |
|  | LBBB | 0.938 | 1.000 | 0.930 | 1.000 | 0.636 | 0.818 (0.669-0.967) | 0.699 (0.427-0.873) | 0.778 |
|  | RBBB | 1.000 | 1.000 | 1.000 | 1.000 | 1.000 | 1.00 (1.00-1.00) | 1.00 (1.00-1.00) | 1.000 |
|  | AVb | 0.906 | 0.692 | 0.961 | 0.925 | 0.818 | 0.871 (0.747-0.996) | 0.598 (0.312-0.818) | 0.750 |

The third of these four open-source datasets consisted of 45,389 ECGs obtained from patients enrolled in the UK Biobank. Here, the model had AUROCs ranging from 0.91 to 0.99 (**Table 9**), thus highlighting the reproducibility of our approach across clinical and non-clinical settings.

Lastly, in 21,784 ECGs from the Germany-based PTB-XL dataset the model had AUROCs of 0.96, 0.95, and 0.91 for AF, ST, and SB, respectively, and 0.97, 0.98 and 0.84 for LBBB, RBBB, and 1dAVb respectively.

## DISCUSSION

We describe the development and external validation of ECG-GPT, a first-of-its-kind AI pipeline that enables the direct generation of automated, complete ECG interpretation statements from images of ECGs in any format. The model performs well against clinician-certified reports across various natural language generation metrics and diagnostic labels spanning a wide array of conduction, rhythm, and structural heart disorders. The model’s scalability is further supported by its robust performance across a range of demographically, temporally, and geographically distinct cohorts, its online deployment, and the capacity to containerize and deploy the model without sharing data in a federated approach.

Our work on format-independent, image-based ECG captioning represents a novel development for vision-text machine learning models. Our model significantly outperforms prior implementations for generating free-text reports from ECG signals, with CIDEr and METEOR scores of 4.69 and 0.75, respectively, compared with scores of 2.55 and 0.27, respectively, reported in a previous study suggesting high textual consistency between original and generated reports.^41^ Machine learning-based multilabel models have been previously developed to simultaneously diagnose large sets of conditions, but these approaches are inherently limited to those labels selected for training and do not capture the full diagnostic range of ECGs.^12,15,42^ Moreover, the utility of such signal models is limited to healthcare systems with the resources to store signal data and incorporate models into the clinical workflow. We demonstrate consistent performance across a range of external validation sets for classifying key rhythm and conduction disorders. Of note, the performance for labels in external validation sets where specific labels were explicitly available matches the performance on those labels in prior published reports. The simplicity of this system, based on images, is that there is inherent interoperability and the absence of a requirement to integrate with ECG machines to extract signals. This approach is particularly advantageous in low-resource regions, where ECGs are currently not stored beyond printing ECG images at the time of acquisition.^43^ This approach also adds convenience and provides access in any venue, including, for example, to emergency medical services providers or in remote locations.

ECG-GPT can be used directly by clinicians at the point of care by uploading ECG images from their phones or as scanned images to a web-based interface, with a demonstration accompanying this study.^44^ This applies anywhere that end-users may still lack access to automated reads or require interpretation before a specialist’s review. The image-based model can also be more easily integrated into repositories of scanned ECGs, the most prevalent and interoperable format for storing and sharing ECGs. Further, ECG-GPT has a unique combination of diagnostic accuracy and range, demonstrating expert-level performance for key conditions while also retaining the capability to generate statements for rare conditions frequently not captured by standard multi-label models. This feature could make ECG-GPT a tool for generating pre-reads and enabling more efficient triage globally in areas with insufficient access to specialists and computerized ECG interpretation algorithms. The study here focuses on the robustness of ECG-GPT in generating accurate interpretations from ECG images, but future prospective studies are warranted to fully assess its integration into clinical workflows in care settings where the tool is used.

Our study has several limitations. First, while the model accurately diagnosed the selected conditions, it is impossible to determine and thus evaluate the performance for the full extent of possible diagnoses the model could output for a given ECG image due to the size and variety of the corpus of diagnosis statements used for model development. However, we report model performance across various rhythm and conduction disorders of varying severity and prevalence, suggesting that ECG-GPT’s performance generalizes to other conditions. Moreover, using the federated approach implemented for external validation, ECG-GPT could be continually fine-tuned to improve performance in individual healthcare systems. This would ensure a reliable pipeline with consistent performance for future ECGs within the specific patient populations in which the model is deployed. Second, it currently only generates interpretation statements in English, which may limit its utility in non-English-speaking settings.

Third, while four different formats were used during model development, we cannot ascertain whether the model generalizes equally well to every other novel ECG image format. However, the model’s performance within the large, native image datasets from MSHS, HMH, and LRH, each consisting of ECG images plotted in configurations distinct from those used in model development, indicates the model can generate accurate interpretation statements for ECG images with variations not seen during training. Finally, though strictly expert-validated diagnosis statements were used to develop the model, these statements are not always completely accurate, limiting the model’s performance. As evidenced by the high diagnostic accuracy of the model in the external set of ECGs manually annotated by two cardiologists, the model may perform better if developed and evaluated in more rigorously validated diagnosis statements.

Furthermore, the current practice of clinicians over-reading the computer-generated reads and correcting them without version control precludes the head-to-head assessment of ECG-GPT against computer-generated reads. Future studies could compare the automated outputs of the model to ECGs labeled by cardiologists without prior computerized interpretation, providing a clearer assessment of the model’s performance relative to human experts. Nevertheless, the higher performance in labels assigned by more than one expert suggests that it likely performs at or above the performance of the current computerized reads at the US health systems, especially for the tested diagnoses.

## CONCLUSIONS

We have developed and extensively validated a novel vision-text transformer capable of generating complete diagnostic statements from ECG images in any lead layout and configuration. Our approach represents a scalable and accessible strategy for generating accurate, expert-level reports from photos of ECGs, enabling accurate ECG interpretation anywhere that an ECG image, paper or digital, can be produced.

## CODE AVAILABILITY

The code for the study is available from the authors for review. Final code will be made available for replication at the time of publication. However, due to intellectual property and IRB stipulations at Yale, as well as patient privacy and data protection regulations, the model weights will not be made publicly available. A completely functional version of the model is publicly hosted at https://www.cards-lab.org/ecg-gpt.

## Data Availability

The development dataset from Yale and the validation datasets from Mount Sinai, Houston Methodist, and Lake Regional Hospital are not publicly available, given the stipulations of the relevant Institutional Review Boards. The external datasets from Brazil, UK Biobank, and PTB-XL are available directly from the respective groups and are outside the purview of the authors.

## Author contributions

RK conceived the study and accessed the data. AK, VS, and RK developed the model. AK, VS, AV, and RK pursued the statistical analysis. AK, VS, and LSD drafted the manuscript. All authors provided feedback regarding the study design and made critical contributions to the manuscript writing. RK supervised the study, procured funding, and is the guarantor.

## ABBREVIATIONS AND ACRONYMS

ECG: Electrocardiography
AI: Artificial Intelligence
YNHH: Yale New Haven Hospital
MSHS: Mount Sinai Health System
HMH: Houston Methodist Hospital
LRH: Lake Regional Hospital
AUROC: Area under receiving operation characteristics
AUPRC: Area under precision recall curve

