## Supplement for "Artificial Intelligence-Based Automated Interpretation of Images of Electrocardiograms: Development and Multinational Validation of ECG-GPT"

**Table S1. Expansion of 100 most common abbreviations and misspellings used for diagnostic statements preprocessing.**

| **Abbreviation or Misspelling** | **Expansion** |
| --- | --- |
| 1d | First degree |
| 1davb | First degree atrioventricular block |
| 1st | First |
| 2d | Second degree |
| 2davb | Second degree atrioventricular block |
| 2nd | Second |
| a-flutter/fibrillation | Atrial fibrillation / atrial flutter |
| abn | Abnormal |
| abnl | Abnormal |
| abnls | Abnormal |
| abnorm | Abnormal |
| abnorma | Abnormal |
| abnorml | Abnormal |
| abnrm | Abnormal |
| abp | Premature atrial contractions |
| af | Atrial fibrillation |
| afib | Atrial fibrillation |
| afib-flutter | Atrial fibrillation / atrial flutter |
| afib/aflutter | Atrial fibrillation / atrial flutter |
| afib/flut | Atrial fibrillation / atrial flutter |
| afib/flutter | Atrial fibrillation / atrial flutter |
| afibrillatiom | Atrial fibrillation |
| afibrillation | Atrial fibrillation |
| aflutter/afib | Atrial fibrillation / atrial flutter |
| alwmi | Anterior and lateral wall myocardial infarction |
| ami | Acute myocardial infarction |
| anteriorlateral | Anterior lateral |
| apc | Premature atrial contractions |
| apcs | Premature atrial contractions |
| asmi | Anteroseptal myocardial infarction |
| aswmi | Anteroseptal myocardial infarction |
| av | Atrioventricular |
| avb | Atrioventricular block |
| awmi | Anteroseptal myocardial infarction |
| ec | Ecg |
| ekg | Ecg |
| elev | Elevation |
| fib | Fibrillation |
| fibrilation | Fibrillation |
| fibrillatiom | Fibrillation |
| first-degree | First degree |
| flutter/afib | Atrial fibrillation / atrial flutter |
| ilbbb | Incomplete left bundle branch block |
| imi | Inferior wall myocardial infarction |
| indeterm | Indeterminate |
| infarct | Infarction |
| inferiorlateral | Inferior lateral |
| irbbb | Incomplete right bundle branch block |
| ivcd | Intraventricular conduction delays |
| iwmi | Inferior wall myocardial infarction |
| laa | Left atrial enlargement |
| lad | Left axis deviation |
| lae | Left atrial enlargement |
| lafb | Left anterior fascicular block |
| lahb | Left anterior fascicular block |
| lbbb | Left bundle branch block |
| lds | Leads |
| livcd | Left intraventricular conduction delays |
| lmi | Left wall myocardial infarction |
| lpfb | Left posterior fascicular block |
| lv | Left ventricular |
| lvh | Left ventricular hypertrophy |
| lwmi | Left wall myocardial infarction |
| mi | Myocardial infarction |
| nl | Normal |
| non-specific | Nonspecific |
| norm | Normal |
| nsc | Neurogenic stress cardiomyopathy |
| nsivcd | Nonspecific interventricular conduction delay |
| nsr | Normal sinus rhythm |
| pac | Premature atrial contractions |
| pacs | Premature atrial contractions |
| pmi | Posterior wall myocardial infarction |
| poor r-wave progression | Poor r wave progression |
| poss | Possible |
| pqt | Prolonged qt interval |
| predom | Predominant |
| prob | Probably |
| prwp | Poor r wave progression |
| pvc | Premature ventricular contractions |
| pvcs | Premature ventricular contractions |
| pwmi | Posterior wall myocardial infarction |
| rbbb | Right bundle branch block |
| repol | Repolarization |
| rivcd | Right intraventricular conduction delays |
| rv | Right ventricular |
| rvh | Right ventricular hypertrophy |
| second-degree | Second degree |
| sr | Sinus rhythm |
| st/t | St-t |
| stemi | St segment elevation myocardial infarction |
| sttab | Significant st-t abnormality |
| suggive | Suggestive |
| sv | Supraventricular |
| svt | Supraventricular tachycardia |
| vcd | Ventricular conduction defect |
| vpb | Premature ventricular contractions |
| vpbs | Premature ventricular contractions |
| vpc | Premature ventricular contractions |
| vpcs | Premature ventricular contractions |

**Table S2.** **Strings used to extract each of the 26 diagnostic labels.** Abbreviations: AF, atrial fibrillation; ST, sinus tachycardia; SB, sinus bradycardia; LBBB, left bundle branch block; RBBB, right bundle branch block; LAFB, left anterior fascicular block; LPFB, left posterior fascicular block; SVT, supraventricular tachycardia; PAC, premature atrial complexes; PVC, premature ventricular complexes; LAE, left atrial enlargement; RAE, right atrial enlargement; LVH, left ventricular hypertrophy; RVH, right ventricular hypertrophy; MI, myocardial infarction; AVb, atrioventricular block; WPW, Wolff-Parkinson-White syndrome; Repol, repolarization. *Acute MI includes ST Elevation MI (STEMI).

| **Condition** | **String Search** |
| --- | --- |
| Sinus Rhythm | Sinus rhythm |
| AF | Atrial fibrillation; afib; auricular fibrillation; fibrillation, atrial; fibrillation, auricular; fibrillations, atrial; fibrillations, auricular |
| Atrial Flutter | Flutter |
| ST | Sinus tachycardia; stach |
| SB | Sinus bradycardia; sbrad |
| Sinus Arrhythmia | Sinus arrhythmia |
| LBBB | Left bundle branch block; lbbb |
| RBBB | Right bundle branch block; rbbb |
| LAFB | Left anterior fascicular block; left anterior hemiblock; lafb; lahb |
| LPFB | Left posterior fascicular block; left posterior hemiblock; lpfb; lphb |
| SVT | Supraventricular tachycardia; atrioventricular reciprocating tachycardia; atrioventricular nodal reentrant tachycardia; svt; atach; atachy; avrt; avnrt |
| PAC | Premature atrial contraction; premature atrial complex; atrial premature complex; atrial premature contraction; premature atrial beat; atrial premature beat; premature supraventricular complex; premature supraventricular contraction; premature supraventricular beat; pac; apc; pab; apb; psvc; psvb; pacs; pac's; apcs; apc's; pabs; pab's; apbs; apb's; psvcs; psvc's; psvbs; psvb's |
| PVC | Premature ventricular contraction; premature ventricular complex; ventricular premature complex; ventricular premature contraction; premature ventricular beat; ventricular premature beat; ventricular couplet; ventricular bigeminy; ventricular trigeminy; ectopic ventricular; pvc; vpc; pvb; vpb; pvcs; pvc's; vpcs; vpc's; pvbs; pvb's; vpbs; vpb's; PV beat; PV complex; VP beat; VP complex |
| LAE | Left atrial abnormality; left atrial enlargement; lae; laa; biatrial |
| RAE | Right atrial abnormality; right atrial enlargement; rae; raa; biatrial |
| LVH | Left ventricular hypertrophy; lvh |
| RVH | Right ventricular hypertrophy; rvh |
| Low Voltage | Low voltage |
| Left Axis Deviation | Left axis deviation; axis deviation, left; lad |
| Acute MI* | Acute mi; ami; stemi; st segment elevation myocardial infarction; st elevation myocardial infarction; st elevation mi; acute myocardial infarct; acute anterolateral infarct; acute anteroseptal infarct; acute inferior infarct; acute lateral infarct; acute posterior infarct; acute septal infarct; acute inferolateral infarct; acute anteroapical infarct; acute apical infarct |
| Lead Reversal | Reversal |
| 1^st^ degree AVb | 1dAVb; prolonged pr interval; first degree atrioventricular block; first degree heart block; first degree AV block; first degree A-V block |
| 2^nd^ degree AVb | 2dAVb; second degree atrioventricular block; second degree heart block; second degree AV block; second degree A-V block; mobitz |
| 3^rd^ degree AVb | 3dAVb; third degree atrioventricular block; third degree heart block; third degree AV block; third degree A-V block; complete block; complete heart block |
| WPW | Wolff-Parkinson-White; Wolff Parkinson White; WPW |
| Repol Abnormality | Repolarization abnormality |

**Table S3. Characteristics of study population.** Data presented mean [SD] for age and number (percent) for other variables. Abbreviations: N, number; MSHS, Mount Sinai Health System, AF, atrial fibrillation; ST, sinus tachycardia; SB, sinus bradycardia; LBBB, left bundle branch block; RBBB, right bundle branch block; AVb, atrioventricular block; LAFB, left anterior fascicular block; LPFB, left posterior fascicular block; SVT, supraventricular tachycardia; PAC, premature atrial complexes; PVC, premature ventricular complexes; LAE, left atrial enlargement; LVH, left ventricular hypertrophy; MI, myocardial infarction.

|  | **Yale Development** | **Yale Test** | **MSHS (Signals)** | **MSHS (PDFs)** | **HMH** |
| --- | --- | --- | --- | --- | --- |
| N | 2888384 | 250589 | 1434455 | 10052 | 62187 |
| N Patients | 601616 | 57860 | 470482 | 6021 | 20087 |
| Male | 1406980 (48.71%) | 124411 (49.65%) | 729795 (50.90%) | 5273 (52.46%) | 27367 (44.01%) |
| Female | 1393583 (48.25%) | 117473 (46.88%) | 704660 (49.10%) | 4689 (46.65%) | 34820 (55.99%) |
| Age | 63.2 (18.0) | 64.4 (17.5) | 61.6 (17.3) | 62.6 (19.3) | 62.0 (11.0) |
| Hispanic | 298490 (10.33%) | 23673 (9.45%) | 56778 (3.96%) | 33 (0.33%) | 7470 (12.01%) |
| White | 1904662 (65.94%) | 170949 (68.22%) | 395443 (27.57%) | 2974 (29.59%) | 40414 (64.99%) |
| Black | 437244 (15.14%) | 31737 (12.66%) | 106305 (7.41%) | 384 (3.82%) | 7919 (12.73%) |
| Asian | 34855 (1.21%) | 2944 (1.17%) | 26898 (1.88%) | 109 (1.08%) | 3898 (6.27%) |
| Other | 213133 (7.37%) | 21286 (8.45%) | 849031 (59.19%) | 6552 (65.18%) | 2486 (4.00%) |
| Sinus Rhythm | 1935574 (67.01%) | 165284 (65.96%) | 956773 (66.67%) | 6350 (63.17%) | 44951 (71.49%) |
| AF | 281961 (9.76%) | 26360 (10.52%) | 104205 (7.26%) | 739 (7.35%) | 1806 (2.87%) |
| Atrial Flutter | 57978 (2.01%) | 5260 (2.1%) | 22434 (1.56%) | 136 (1.35%) | 341 (0.54%) |
| ST | 265640 (9.2%) | 20293 (8.1%) | 155167 (10.81%) | 1018 (10.13%) | 2148 (3.42%) |
| SB | 126757 (4.39%) | 13495 (5.39%) | 162114 (11.3%) | 1114 (11.08%) | 11346 (18.05%) |
| Sinus Arrythmia | 61090 (2.12%) | 6209 (2.48%) | 64267 (4.48%) | 473 (4.71%) | 2931 (4.66%) |
| LBBB | 94298 (3.26%) | 8581 (3.42%) | 35793 (2.49%) | 265 (2.64%) | 718 (1.14%) |
| RBBB | 195474 (6.77%) | 17994 (7.18%) | 96502 (6.72%) | 691 (6.87%) | 2494 (3.97%) |
| LAFB | 155593 (5.39%) | 13477 (5.38%) | 52363 (3.65%) | 371 (3.69%) | 1361 (2.16%) |
| LPFB | 15293 (0.53%) | 1343 (0.54%) | 5917 (0.41%) | 32 (0.32%) | 185 (0.29%) |
| SVT | 11478 (0.4%) | 1038 (0.41%) | 4663 (0.32%) | 44 (0.44%) | 133 (0.21%) |
| PAC | 153698 (5.32%) | 14699 (5.87%) | 83985 (5.85%) | 642 (6.39%) | 1970 (3.13%) |
| PVC | 201456 (6.97%) | 18349 (7.32%) | 90400 (6.3%) | 675 (6.72%) | 2333 (3.71%) |
| LAE | 359747 (12.45%) | 29004 (11.57%) | 138457 (9.65%) | 847 (8.43%) | 2819 (4.48%) |
| RAE | 15075 (0.52%) | 1091 (0.44%) | 9260 (0.65%) | 65 (0.65%) | 437 (0.7%) |
| LVH | 342340 (11.85%) | 27792 (11.09%) | 182728 (12.73%) | 1369 (13.62%) | 3476 (5.53%) |
| RVH | 36863 (1.28%) | 2910 (1.16%) | 12764 (0.89%) | 45 (0.45%) | 163 (0.26%) |
| Low Voltage | 235069 (8.14%) | 21802 (8.7%) | 82277 (5.73%) | 548 (5.45%) | 3897 (6.2%) |
| Left Axis Deviation | 256740 (8.89%) | 22865 (9.12%) | 134860 (9.4%) | 1028 (10.23%) | 3505 (5.57%) |
| Acute MI* | 20003 (0.69%) | 1624 (0.65%) | 13650 (0.95%) | 60 (0.6%) | 115 (0.18%) |
| Lead Reversal | 4236 (0.15%) | 425 (0.17%) | 3616 (0.25%) | 40 (0.4%) | 101 (0.16%) |
| 1^st^ Degree AVb | 211836 (7.33%) | 18501 (7.38%) | 84083 (5.86%) | 694 (6.9%) | 2939 (4.67%) |
| 2^nd^ Degree AVb | 3443 (0.12%) | 353 (0.14%) | 2797 (0.19%) | 20 (0.2%) | 51 (0.08%) |
| 3^rd^ Degree AVb | 2988 (0.1%) | 238 (0.09%) | 1219 (0.08%) | 17 (0.17%) | 28 (0.04%) |
| WPW | 1137 (0.04%) | 133 (0.05%) | 983 (0.07%) | 7 (0.07%) | 23 (0.04%) |
| Repol Abnormality | 227309 (7.87%) | 18193 (7.26%) | 59788 (4.17%) | 366 (3.64%) | 867 (1.38%) |

**Table S4. Performance of model on test images across natural language generation metrics in held-out test set.** Abbreviations: NLG, natural language generation; ROUGE, Recall Oriented Understudy of Gisting Evaluation; BLEU, BiLingual Evaluation Understudy; METEOR, Metric for Evaluation of Translation with Explicit ORdering; CIDEr, Consensus-based Image Description Evaluation.

| **NLG Metric** | **Performance** |
| --- | --- |
| ROUGE-1 | 0.663 |
| ROUGE-L | 0.655 |
| BLEU-1 | 0.595 |
| BLEU-2 | 0.529 |
| BLEU-3 | 0.465 |
| BLEU-4 | 0.419 |
| METEOR | 0.693 |
| CIDEr | 3.321 |

**Table S5.** **Comparison of ECG-GPT model performance to two previously published CNN models on ECG signals.** Abbreviations: AF, atrial fibrillation; ST, sinus tachycardia; SB, sinus bradycardia; LBBB, left bundle branch block; RBBB, right bundle branch block; LAFB, left anterior fascicular block; LPFB, left posterior fascicular block; SVT, supraventricular tachycardia; PAC, premature atrial complexes; PVC, premature ventricular complexes; LAE, left atrial enlargement; RAE, right atrial enlargement; LVH, left ventricular hypertrophy; RVH, right ventricular hypertrophy; MI, myocardial infarction; AVb, atrioventricular block; WPW, Wolff-Parkinson-White syndrome; Repol, repolarization. *Acute MI includes ST Elevation MI (STEMI).

|  | **ECG-GPT** | | | **Hughes et al** | | | **Kashou et al**** | | |
| --- | --- | --- | --- | --- | --- | --- | --- | --- | --- |
|  | **Specificity** | **Sensitivity** | **F1** | **Specificity** | **Sensitivity** | **F1*** | **Specificity** | **Sensitivity** | **F1** |
| **Sinus Rhythm** | 0.944 | 0.954 | 0.962 | 0.94 | 0.75 | 0.849 | 0.957 | 0.877 | 0.853 |
| **AFIB** | 0.983 | 0.862 | 0.858 | 0.99 | 0.8 | 0.847 | 0.694 | 1 | 0.38 |
| **Atrial Flutter** | 0.970 | 0.868 | 0.529 | 0.99 | 0.667 | 0.75 | 0.704 | 1 | 0.118 |
| **ST** | 0.994 | 0.912 | 0.924 | NA | NA | NA | 0.643 | 1 | 0.302 |
| **SB** | 0.995 | 0.867 | 0.885 | NA | NA | NA | 0.968 | 0.994 | 0.911 |
| **Sinus Arrhythmia** | 0.964 | 0.922 | 0.553 | NA | NA | NA | 0.679 | 0.995 | 0.221 |
| **LBBB** | 0.995 | 0.855 | 0.854 | 0.99 | 0.769 | 0.87 | 0.912 | 1 | 0.437 |
| **RBBB** | 0.993 | 0.935 | 0.922 | 0.95 | 1 | 0.941 | 0.932 | 1 | 0.649 |
| **LAFB** | 0.980 | 0.808 | 0.751 | 0.983 | 0.629 | 0.756 | 0.844 | 0.979 | 0.208 |
| **LPFB** | 0.988 | 0.861 | 0.412 | 0.993 | 0.435 | 0.656 | 0.939 | 0.999 | 0.353 |
| **SVT** | 0.996 | 0.698 | 0.508 | 0.984 | 0.5 | 0.696 | 0.875 | 0.998 | 0.043 |
| **PAC** | 0.989 | 0.616 | 0.685 | 0.98 | 0.679 | 0.759 | 0.584 | 0.999 | 0.248 |
| **PVC** | 0.983 | 0.848 | 0.823 | 0.976 | 0.719 | 0.786 | 0.548 | 1 | 0.228 |
| **LAE** | 0.956 | 0.706 | 0.692 | 0.947 | 0.324 | 0.432 | 0.71 | 0.987 | 0.214 |
| **RAE** | 0.990 | 0.764 | 0.377 | 0.975 | 0.889 | 0.875 | 0.628 | 0.998 | 0.024 |
| **LVH** | 0.960 | 0.864 | 0.791 | 0.947 | 0.875 | 0.7 | 0.936 | 0.964 | 0.695 |
| **RVH** | 0.928 | 0.822 | 0.206 | 0.975 | 0.818 | 0.706 | 0.89 | 0.998 | 0.045 |
| **Low Voltage** | 0.963 | 0.762 | 0.708 | 0.938 | 1 | 0.75 | 0.92 | 0.956 | 0.553 |
| **Left Axis Deviation** | 0.959 | 0.753 | 0.697 | 0.964 | 0.574 | 0.8 | 0.793 | 0.989 | 0.05 |
| **Acute MI** | 0.986 | 0.711 | 0.375 | NA | NA | NA | NA | NA | NA |
| **Lead Reversal** | 0.997 | 0.718 | 0.447 | 0.994 | 0.917 | 0.917 | NA | NA | NA |
| **1dAVb** | 0.978 | 0.787 | 0.764 | 0.975 | 0.553 | 0.679 | 0.958 | 0.937 | 0.826 |
| **2dAVb** | 0.999 | 0.643 | 0.477 | 0.994 | 0.889 | 0.941 | 0.879 | 0.996 | 0.025 |
| **3dAVb** | 0.998 | 0.693 | 0.378 | NA | NA | NA | 0.961 | 0.985 | 0.068 |
| **WPW** | 1.000 | 0.624 | 0.519 | 0.991 | 0.75 | 0.8 | 0.925 | 0.985 | 0.01 |
| **Repol Abnormality** | 0.951 | 0.661 | 0.579 | NA | NA | NA | 0.573 | 0.994 | 0.071 |

* The authors presented the maximum possible F1 score, rather than the F1 score at the cutoffs the specificities and sensitivities were reported.

** The authors presented specificities, sensitivities, and F1 scores at 2 different cutoffs, we have reported the values that corresponded to the higher F1 score for each condition

**Table S6. Characteristics of additional external validation datasets.** Data presented mean [SD] for age and number (percent) for other variables. Abbreviations: LRH, Lake Regional Hospital, AF, atrial fibrillation; ST, sinus tachycardia; SB, sinus bradycardia; LBBB, left bundle branch block; RBBB, right bundle branch block; 1dAVb, first-degree atrioventricular block.

|  | **Cardiologist Validated CODE15** | **CODE15** | **UK Biobank** | **PTB-XL** | **LRH** |
| --- | --- | --- | --- | --- | --- |
| Number | 827 | 2228236 | 45350 | 21785 | 64 |
| Female | 506 (61.2%) | 1343866 (60.3%) | 23359 (51.5%) | 10442 (47.9%) | 31 (48.4%) |
| Age | 54.9 (16.5) | 53.6 (17.4) | 63.9 (7.8) | 59.8 (17.0) | 67.2 (16.7) |
| AF | 13 (1.6%) | 39662 (1.78%) | 662 (1.46%) | 1507 (6.9%) | 13 (20.31%) |
| ST | 37 (4.5%) | 48296 (2.17%) | 92 (0.2%) | 825 (3.8%) | 10 (15.63%) |
| SB | 16 (1.9%) | 35441 (1.59%) | 4273 (9.42%) | 637 (2.9%) | 10 (15.63%) |
| LBBB | 30 (3.6%) | 34677 (1.56%) | 397 (0.88%) | 522 (2.4%) | 11 (17.19%) |
| RBBB | 34 (4.1%) | 61551 (2.76%) | 891 (1.96%) | 541 (2.5%) | 11 (17.19%) |
| 1dAVb | 28 (3.4%) | 34446 (1.55%) | 2503 (5.52%) | 793 (3.6%) | 11 (17.19%) |

**Figure S1. Flow chart of study cohort and analysis.** Abbreviations: ECG, electrocardiogram.

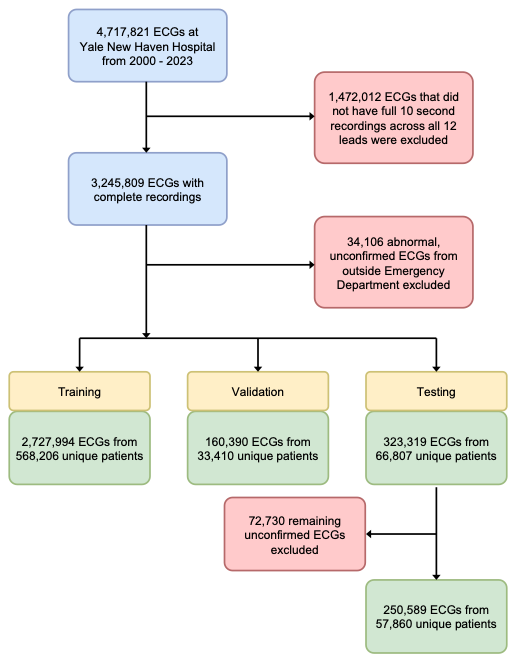

**Figure S2: Example of real-world image that met threshold A) before and B) after brightness and contrast correction**

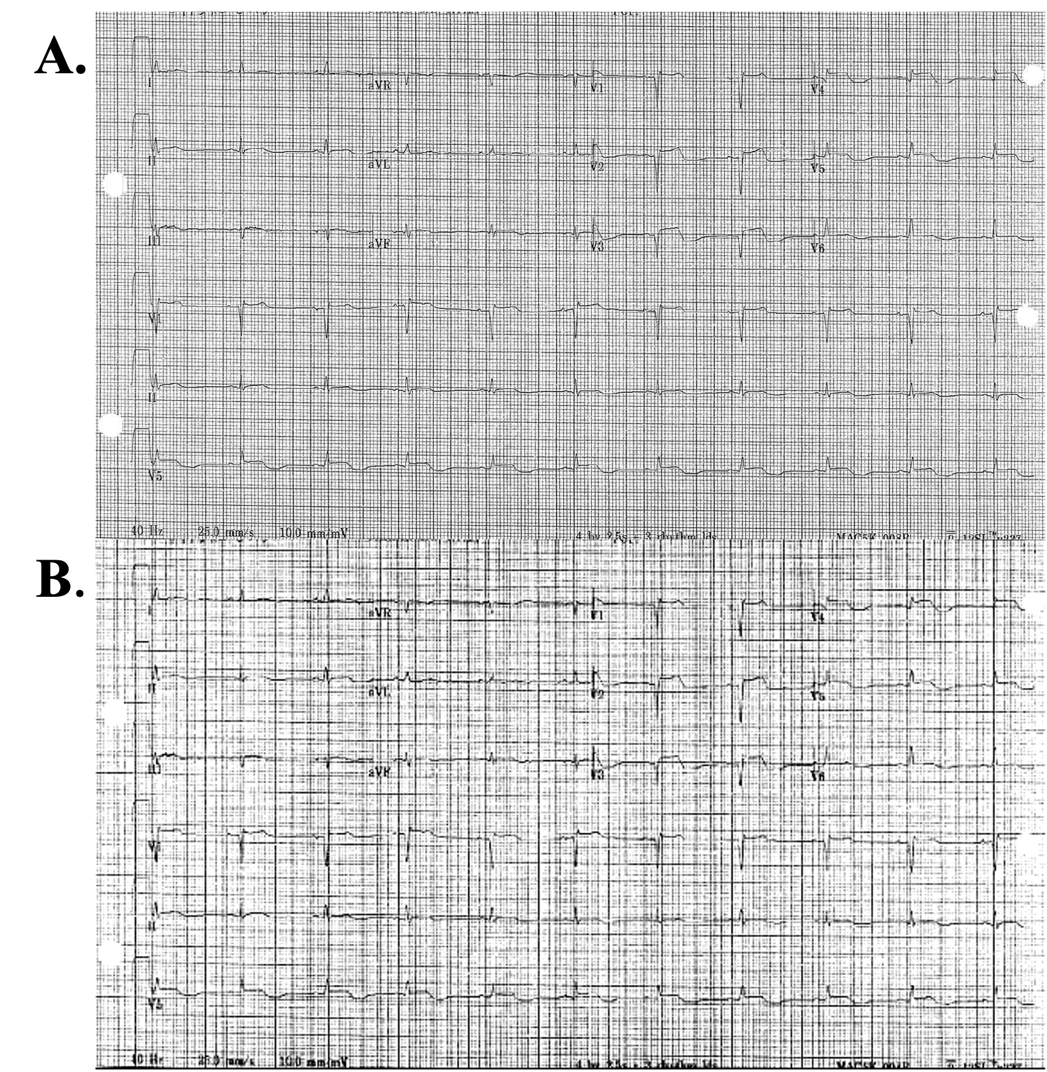

**Figure S3: Examples of synthetic (A and B) and real world C) images that failed the quality control threshold.**

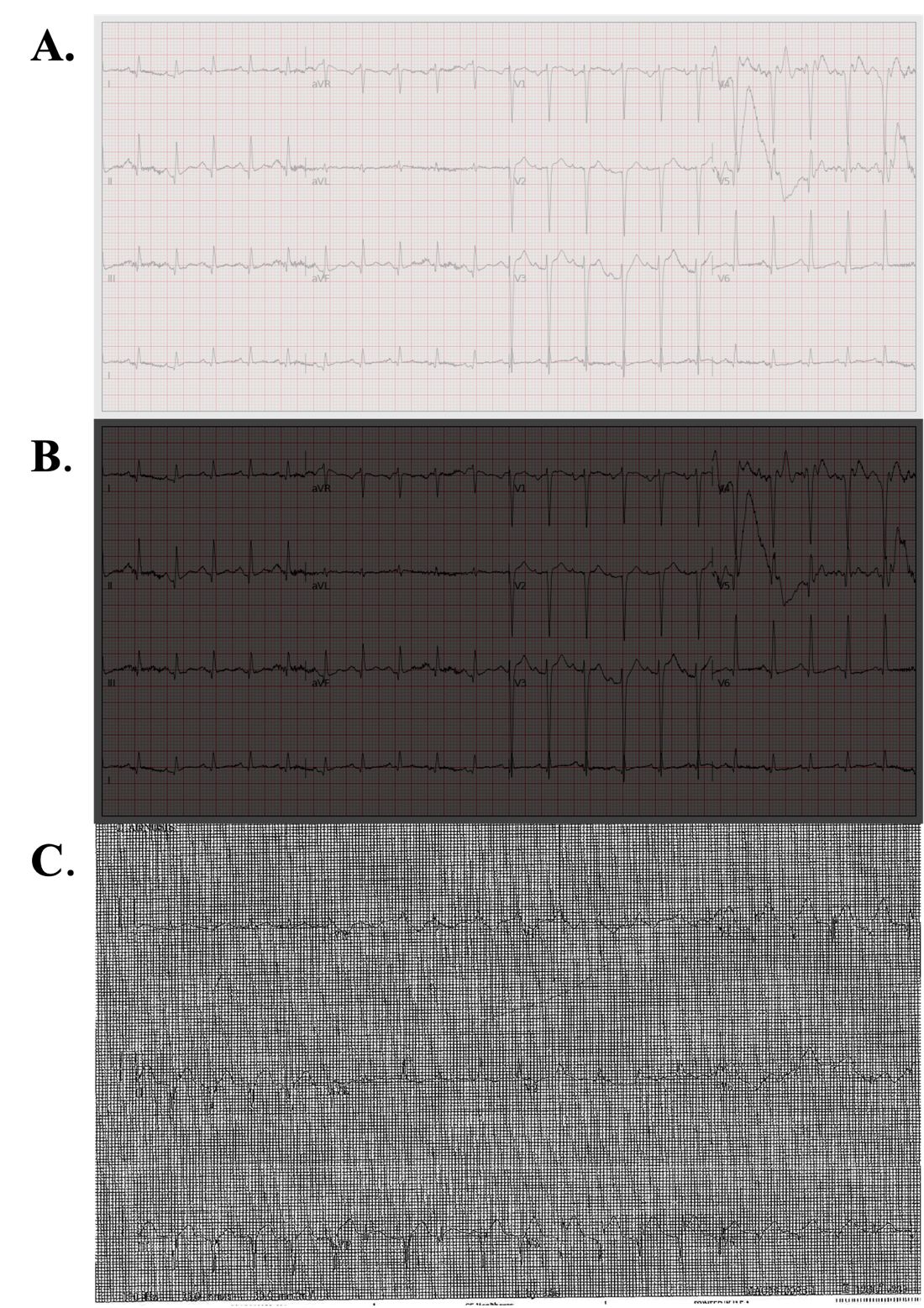

**Figure S4. Preprocessing of Cardiologist Diagnosis Statements for Model Development and Evaluation.** Abbreviations: ECG, electrocardiogram; LVH, left ventricular hypertrophy.

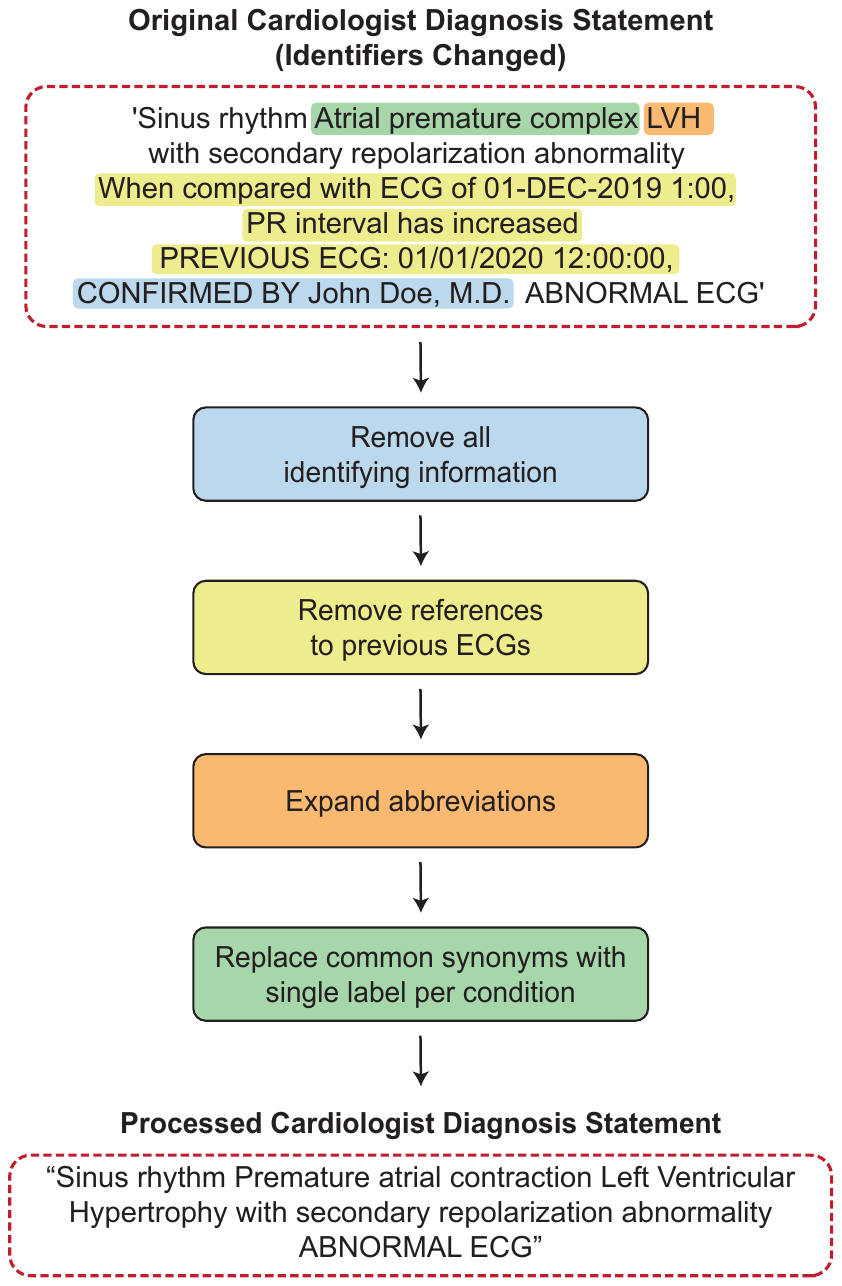
